# Impact of autumn 2023 and 2024 COVID-19 vaccination in preventing COVID-19 related hospitalisations and deaths in seven EU/EEA countries: a VEBIS-EHR network study

**DOI:** 10.64898/2026.07.10.26357728

**Authors:** Yohann Mansiaux, Alexandre Blake, Nathalie Nicolay, James Humphreys, Toon Braeye, Izaak Van Evercooren, Christian Holm Hansen, Ida Rask Moustsen-Helms, Daniele Petrone, Alberto Mateo-Urdiales, Iván Martínez-Baz, Jesús Castilla, Ausenda Machado, Patricia Soares, Rickard Ljung, Nicklas Pihlstrom, Hinta Meijerink, Anthony Nardone, Esther Kissling, Sabrina Bacci, Susana Monge, Baltazar Nunes, VEBIS-EHR working group

## Abstract

**Background:** Within the VEBIS-EHR project, monthly vaccine effectiveness (VE) of COVID-19 vaccines is routinely estimated across EU/EEA countries. While VE quantifies direct protection, it does not capture the overall population benefit of vaccination campaigns in terms of severe outcomes prevented.

**Aim:** To estimate the impact of the 2023 and 2024 autumn COVID-19 vaccination campaigns in adults aged ≥65 years.

**Methods:** We conducted a retrospective cohort study using electronic health records data from Belgium, Denmark, Italy, Navarre (Spain), Portugal, Norway and Sweden. Weekly numbers of averted COVID-19-related hospitalisations and deaths during the 12 months following each campaign were estimated using observed COVID-19-related events, vaccine coverage (VC) and interpolated weekly VE.

**Results:** Across participating countries/regions, among adults aged ≥65 years, the 2023 autumn vaccination campaign averted approximately 6,200 hospitalisations (prevented fraction [PF] 10%) compared with 2,200 (PF 12%) in 2024. Among those aged ≥80 years, the number of averted COVID-19-related deaths was 811 (PF 13%) for the 2023 campaign and 156 (PF 12%) for the 2024 campaign. Impact varied across countries, reflecting differences in VC, vaccination timing and outcome occurrence.

**Conclusion:** The 2023 and 2024 autumn vaccination campaigns resulted in substantially different numbers of averted COVID-19-related hospitalisations and deaths among older adults, with fewer events averted in 2024. These findings highlight that the impact of vaccination programmes depends not only on VC and VE but also on alignment between vaccination timing and periods of increased viral circulation.

## Introduction

Measuring the overall effect of a vaccination programme is essential to inform and evaluate vaccination campaigns. Within the Vaccine Effectiveness, Burden and Impact Studies using Electronic Health Records project (VEBIS-EHR), we routinely estimate vaccine effectiveness (VE) of COVID-19 vaccines administered as part of the annual autumn vaccination campaigns in participating EU/EEA countries (1–4). These monthly estimates provide timely evidence on the level of direct protection conferred by vaccination during each COVID-19 wave, including specific periods of interest such as Omicron sub-lineage predominance or periods of high incidence (e.g. summer 2024) (5–7). However, VE estimates do not quantify the population level benefit of vaccination, in terms of the number of severe events averted.

Previous studies estimating the impact of COVID-19 vaccination programmes in EU/EEA countries or regions (8–12) focused on evaluating the impact of primary series vaccination up to the second booster dose, conducted between 2020 and 2022, during the periods when Alpha, Delta, and early Omicron variants were circulating. Since then, COVID-19 vaccines have been adapted to better match circulating Omicron sublineages, with updated antigenic compositions introduced from 2023 onwards (13). To our knowledge, only two studies have measured the impact of the more recently adapted COVID-19 vaccines (in the 2023-2024 season), in Italy and Navarre (Spain) (14,15). However, evidence on the population-level impact of these vaccination campaigns across multiple European settings remains limited, particularly in the post-pandemic context.

This study aimed to estimate the impact of the 2023 and 2024 autumn COVID-19 vaccination campaigns on COVID-19-related hospitalisations and deaths among adults aged 65 years and over across seven EU/EEA countries, during the 12 months following two subsequent vaccination campaign (from 2023-W40 to 2025-W39).

## Methods

### Study design, population and study period

We implemented a retrospective cohort study to obtain estimates for the impact of the 2023 and 2024 autumn COVID-19 vaccination campaigns using population EHR data obtained from seven study sites within VEBIS-EHR network (Belgium, Denmark, Italy, Navarre – Spain, Portugal, Norway and Sweden) (the study protocol can be found online (16)).

The study population comprised community-dwelling individuals aged 65 years and older, identified from national/regional EHR databases. Participants were required to be eligible for the 2023 and 2024 autumn COVID-19 vaccination (see Table S1 for vaccination campaign dates), including belonging to an age group for whom seasonal vaccination was recommended at each site or country. More details on the definition of the study population can be found in the VEBIS-EHR scientific protocols (17,18). The analysis was stratified by age group: individuals aged 65 –79 years and those aged ≥80 years old. The study period was from weeks 2023-W40 (from 2 October 2023) to 2024-W39 (to 29 September 2024) for the 2023–2024 study period and from 2024-W40 (from 30 September 2024) to 2025-W39 (to 28 September 2025) for the 2024–2025 study period.

### Outcomes, vaccine coverage and vaccine effectiveness

The outcomes of interest were hospital admission due to COVID-19 and COVID-19-related death. Hospitalisation was defined as a hospital admission with COVID-19 as the main diagnosis in the discharge record or with admission criteria compatible with a severe acute respiratory infection and a laboratory-confirmed SARS-CoV-2 infection between 14 days before and 24 hours after admission. A COVID-19-related death was defined as a death for which COVID-19 was recorded as the main or underlying cause of death or, if cause of death is not available, laboratory-confirmed SARS-CoV-2 infection within 30 days before death. We used the weekly number of COVID-19-related hospitalisations and deaths in the eligible population. For Belgium, COVID-19-related hospitalisations were extracted from the SARI surveillance system as this data source was more exhaustive (see supplementary material – section 2).

Weekly vaccine coverage was calculated in each study site. For each week t, the proportion of individuals vaccinated was estimated by 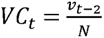, where *N* is the number of individuals eligible for vaccination at the start of the study period and *v*_*t*-2_ is the cumulative number of vaccinated individuals up to week t - 2 (2 weeks prior, to allow individuals to gain immunity after vaccination) among the individuals eligible for vaccination at the start of the study period. Participants vaccinated before the start of the study were excluded from the numerator and denominator. Therefore, the vaccination impact was estimated from the third week of the study period onward: from 2023-W42 for the 2023 autumn vaccination campaign, and from 2024-W42 for the 2024 autumn vaccination campaign.

Cox regression with calendar time as the underlying time scale was used to estimate the confounder-adjusted hazard ratios (aHR) for vaccination status association with the outcomes, at study site level. Models included as confounders 5-year age group, sex, region within each country, comorbidity status as defined at study site level (none, medium-risk and high-risk/immunocompromising – Table S2), and number of previous COVID-19 booster doses. Each site estimated monthly vaccination status aHR using 8-week rolling periods starting on day 1 of each month (Table S3). VE estimates were obtained by the formula VE = (1 – pooled aHR) x100, where pooled aHR was calculated by the Paule-Mandel random-effects meta-analysis method (19). Details on the methods used to estimate the VE at study site level and pooled level can be found in the VEBIS-EHR master protocols (17,18).

The 8-week VE estimates were interpolated to estimate weekly VE during the study period. They were modelled by either an exponential or a logistic decay function, with the most parcimonious model selected using the Bayesian information criterion (BIC). 8-week VE estimates and additional details about the interpolation process are provided in supplementary material. In a sensitivity analysis we assumed constant VE values for each month, using VE sampled from the 8-week estimates (see supplementary material – section 3).

### Impact of the vaccination programmes: number of averted events

We applied an adapted version of methods used to estimate influenza vaccination impact (20,21). Briefly, the number of weekly averted events (NAE) by the 2023 and 2024 vaccination campaign at study site was estimated by: 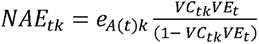 where *e*_*A(t)k*_ is number of events for the population of study site k during week t, *Vc_tk_* is the vaccine coverage for the population of study site k during week t, *VE_k_* is the pooled VE estimate during week t. The expected number of weekly events without the vaccination programme (NEXP) at each study site was estimated as *NEXP_tk_* = *e_A(t)k_* + *NAE_tk_*. Using NAE and NEXP, the weekly proportion of events averted by vaccination, or the prevented fraction (PF), at each study site was estimated as 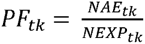. All-study-period estimates of the number of observed events, NAE and PF were obtained by summing study site *e_AK_* and estimates of *NAE_k_* and *NEXP_k_*. To account for the uncertainty around the VE estimates, we implemented a resampling procedure repeated 1,000 times (see supplementary material – section 3). The resulting sets of weekly VE estimates were used to produce a 95% quantile prediction interval (PI).

This uncertainty around the weekly VE was then used in the NAE estimates.

Statistical analysis was performed using R4.2.2. The packages “vaccinationimpact” (22) and “VEinterpolation” (available upon request), developed for this study, were used.

## Results

### COVID-19 hospitalisations, deaths and vaccine coverage

Across both study periods, the study population comprised approximately 14 million adults aged 65–79 years and 6 million aged ≥80 years. Vaccine coverage varied substantially across countries, with lower uptake in Italy and higher uptake in Nordic countries.

In the 2023–2024 study period, the maximum (*i.e.* end-of- season) vaccine coverage ranged from 10 to 81% among adults aged 65–79 years and from 14% to 85% among those aged ≥80 years; in the 2024–2025 study period, it ranged from 4% to 79% and from 6% to 83%, respectively. Coverage was consistently higher in adults aged ≥80 years and lower across all countries in the 2024 autumn campaign than in 2023 autumn vaccination campaign (Table 1). In both campaigns, vaccine coverage increased steadily before reaching a plateau around December–January (Figure 1).

**Figure 1:**
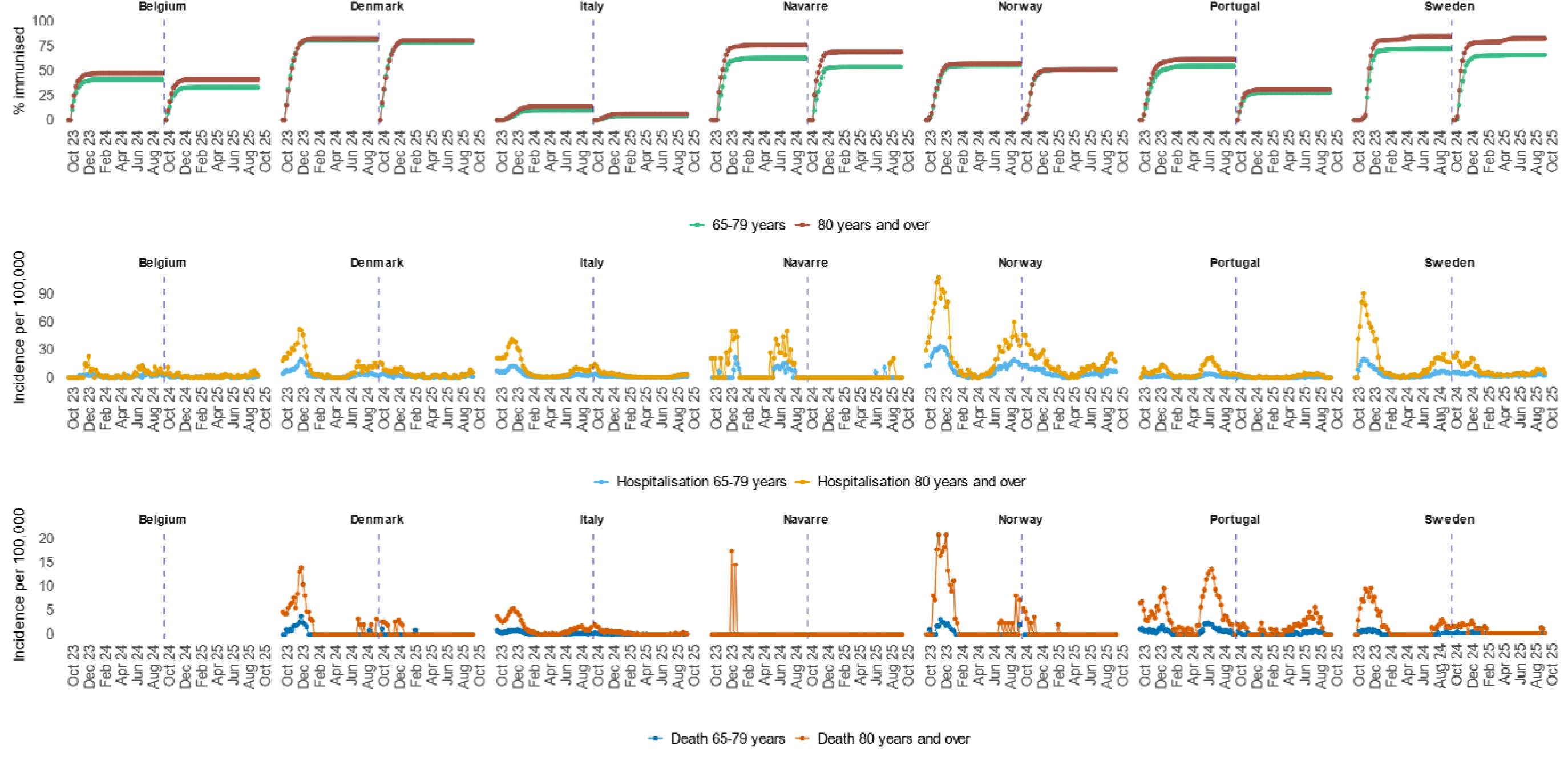
Vaccination coverage, hospital admission due to COVID-19 and COVID-19-related death incidence, October 2023–September 2024 and October 2024–September 2025, VEBIS-EHR, Europe (the dashed line corresponds to the separation between the two study periods)

**Table 1:** Description of the populations included to estimate impact of 2023 and 2024 autumn COVID-19 vaccination campaigns against hospital admissions due to COVID-19 and COVID-19 -related deaths, October 2023–September 2024 and October 2024–September 2025, VEBIS-EHR, Europe.

| Study site by age group | Population included | Cumulative vaccine coverage | Number of hospitalisations | Rate of hospitalisations (per 100,000 population) | Number of deaths | Rate of deaths (per 100,000 population) | Population included | Cumulative vaccine coverage | Number of hospitalisations | Rate of hospitalisations (per 100,000 population) | Number of deaths | Rate of deaths (per 100,000 population) |
| --- | --- | --- | --- | --- | --- | --- | --- | --- | --- | --- | --- | --- |
| 2023–2024 study period: October 2023–September 2024 |  |  |  |  |  |  | 2024–2025 study period: October 2024–September 2025 |  |  |  |  |  |
| **65-79 years** |  |  |  |  |  |  |  |  |  |  |  |  |
| Belgium | 1,438,934 | 40.8% | 1,160 | 81 | .. | .. | 1,605,531 | 33.1% | 543 | 34 | .. | .. |
| Denmark | 698,868 | 81.0% | 1,396 | 200 | 148 | 21 | 642,569 | 79.1% | 399 | 62 | 12 | 2 |
| Italy | 8,578,078 | 10.0% | 14,426 | 168 | 1,055 | 12 | 8,630,464 | 4.3% | 3,550 | 41 | 204 | 2 |
| Navarre | 84,944 | 62.7% | 145 | 171 | 0 | 0 | 85,863 | 53.8% | 14 | 16 | 0 | 0 |
| Norway | 699,035 | 55.0% | 4,058 | 581 | 144 | 21 | 662,164 | 50.9% | 1,682 | 254 | 0 | 0 |
| Portugal | 1,536,055 | 54.5% | 927 | 60 | 538 | 35 | 1,656,318 | 27.9% | 195 | 12 | 128 | 8 |
| Sweden | 1,386,375 | 72.2% | 3,163 | 228 | 126 | 9 | 1,400,449 | 66.0% | 1,329 | 95 | 20 | 1 |
| **≥80 years** |  |  |  |  |  |  |  |  |  |  |  |  |
| Belgium | 498,239 | 47.3% | 1,022 | 205 | .. | .. | 594,063 | 41.2% | 668 | 112 | .. | .. |
| Denmark | 261,634 | 82.2% | 1,678 | 641 | 325 | 124 | 277,339 | 80.4% | 579 | 209 | 61 | 22 |
| Italy | 4,077,709 | 13.8% | 22,730 | 557 | 3,071 | 75 | 4,139,413 | 6.2% | 5,638 | 136 | 657 | 16 |
| Navarre | 34,460 | 75.9% | 234 | 679 | 11 | 32 | 34,983 | 68.8% | 18 | 51 | 0 | 0 |
| Norway | 225,874 | 57.1% | 3,671 | 1,625 | 455 | 201 | 259,043 | 51.1% | 1,960 | 757 | 64 | 25 |
| Portugal | 665,778 | 61.4% | 2,216 | 333 | 1,561 | 234 | 655,335 | 30.7% | 468 | 71 | 453 | 69 |
| Sweden | 531,451 | 84.7% | 5,105 | 961 | 570 | 107 | 586,243 | 82.5% | 2,250 | 384 | 170 | 29 |

Hospitalisation and death rates varied across countries and age groups, with higher rates among adults aged ≥80 years. For both age groups, rates were higher in 2023–2024 than in 2024–2025, with the highest levels observed in Norway (hospitalisations) and Portugal (deaths) (Table 1).

All sites reported increased rates in late 2023–early 2024, followed by a secondary rise in spring–summer 2024 (Figure 1). The 2024–2025 period began with high levels that declined until February–March 2025, followed by a smaller increase in spring–summer 2025. Across sites, autumn–winter peaks occurred before maximum vaccine coverage was reached.

### Weekly vaccine effectiveness

For both study periods, both age groups and both outcomes (except the death outcome in those aged 65–79 years) the logistic decay (vs the exponential) models were the best fit with lower BIC (Table S4). These models were used to estimate weekly VE. For VE against COVID-19-related deaths, missing data (because the number of events was too low for reliable VE estimation) over an 8-week period in the 65–79 years age group did not allow the interpolation and subsequent impact estimation for either study period.

In both seasons VE against both outcomes were higher in the first weeks after the start of the vaccination campaigns (VE varied between 60 and 70%) decreasing with calendar time (below 50% after the first weeks of 2024 and 2025, for the 2023–2024 and 2024–2025 autumn vaccination campaigns, respectively) to residual or null protection at the end of the study periods (Figure 2, Table S5 and S6, Figure S1 to Figure S6).

**Figure 2:**
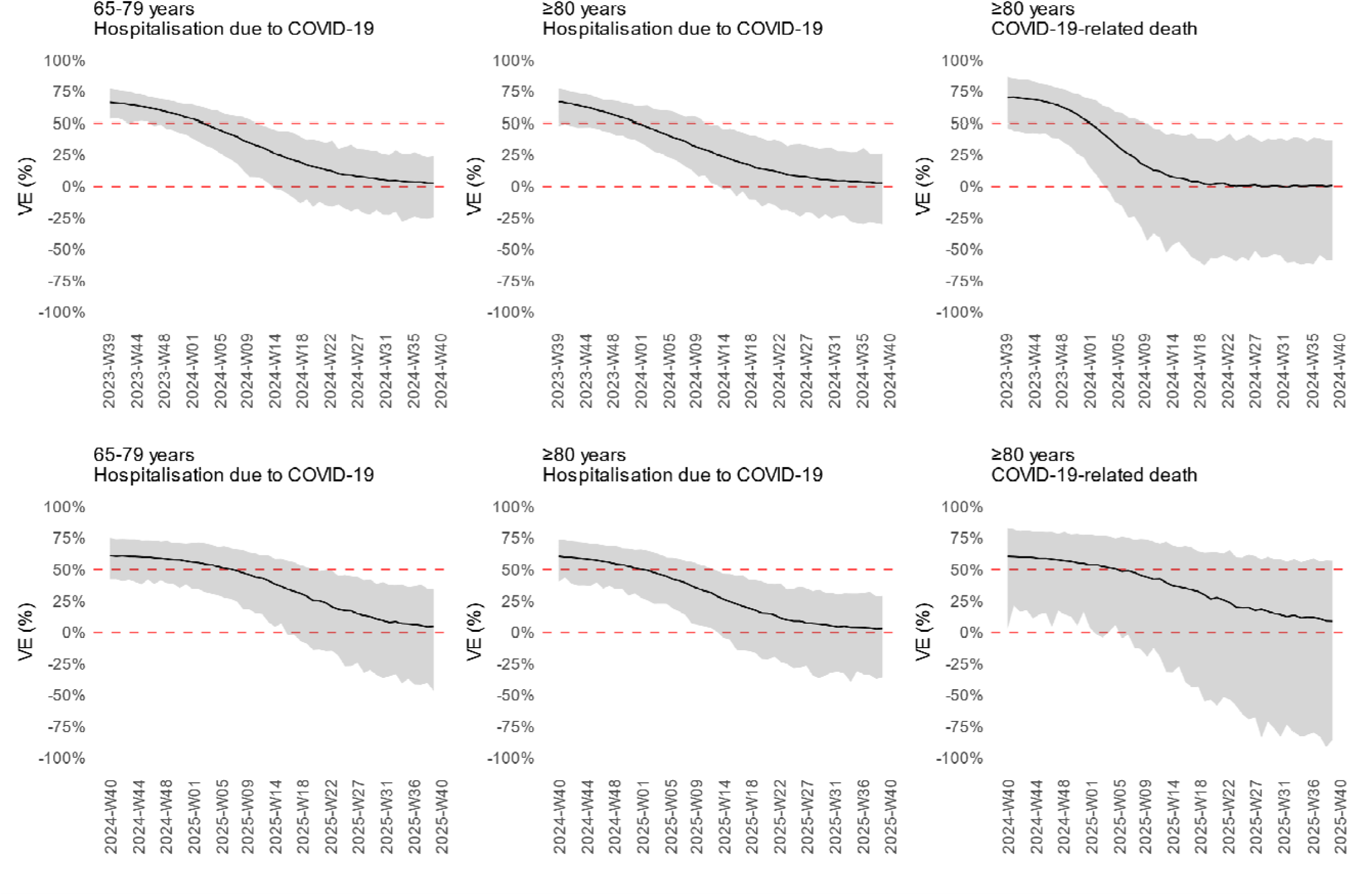
Interpolated weekly vaccine effectiveness against COVID-19, October 2023–September 2024 and October 2024–September 2025, VEBIS-EHR, Europe (the line corresponds to the median and the ribbon to the 95% prediction interval across 1000 repetitions

### Impact of the 2023 and 2024 autumn vaccination campaigns

During the 2023–2024 study period and across all VEBIS-EHR study sites we estimated 2,542 (95% PI: 2,145–2,978) and 3,629 (3,089–4,432) hospitalisations averted by the vaccination, respectively in the 65–79 and ≥80 years age groups (median PF were 10% and 9%). The rate of COVID-19 hospitalisations averted per 100,000 individuals ranged from 3 in Italy to 108 in Norway, and from 13 in Italy to 309 in Norway among those aged 65–79 and ≥80 years, respectively (Table 2).

**Table 2:** Estimates of 2023 and 2024 autumn COVID-19 vaccination campaign impact against hospital admissions due to COVID-19 and COVID-19-related deaths, among individuals aged 65–79 years old and 80 years or older, October 2023–September 2024 and October 2024–September 2025, VEBIS-EHR, Europe.

| Indicator |  | Pooled | Belgium | Denmark | Italy | Navarra | Norway | Portugal | Sweden |
| --- | --- | --- | --- | --- | --- | --- | --- | --- | --- |
| Hospital admission due to COVID-19 |  |  |  |  |  |  |  |  |  |
| **65–79 years old** |  |  |  |  |  |  |  |  |  |
| Events observed included in impact estimation | 2023-2024 | 23,962 | 1,160 | 1,324 | 13,369 | 145 | 3,886 | 915 | 3,163 |
|  | 2024-2025 | 6,641 | 434 | 361 | 2,911 | 14 | 1,515 | 178 | 1,228 |
| NAE | 2023-2024 | 2,542<br>(2,145 - 2,978) | 137<br>(104 - 173) | 684<br>(573 - 795) | 253<br>(211 - 296) | 27<br>(18 - 38) | 755<br>(643 - 875) | 121<br>(85 - 168) | 566<br>(470 - 670) |
|  | 2024-2025 | 859<br>(686 - 1,033) | 48<br>(34 - 61) | 161<br>(129 - 196) | 32<br>(25 - 37) | 1<br>(-1 - 3) | 271<br>(216 - 331) | 8<br>(2 - 15) | 337<br>(270 - 404) |
| NAE / pop. size (per 100,000) | 2023-2024 | 18<br>(15 - 21) | 10<br>(7 - 12) | 98<br>(82 - 114) | 3<br>(2 - 3) | 32<br>(21 - 45) | 108<br>(92 - 125) | 8<br>(6 - 11) | 41<br>(34 - 48) |
|  | 2024-2025 | 6<br>(5 - 7) | 3<br>(2 - 4) | 25<br>(20 - 30) | 0<br>(0 - 0) | 1<br>(-1 - 4) | 41<br>(33 - 50) | 0<br>(0 - 1) | 24<br>(19 - 29) |
| PF (%) | 2023-2024 | 10<br>(8 - 11) | 11<br>(8 - 13) | 34<br>(30 - 38) | 2<br>(2 - 2) | 16<br>(11 - 21) | 16<br>(14 - 18) | 12<br>(9 - 16) | 15<br>(13 - 17) |
|  | 2024-2025 | 11<br>(9 - 13) | 10<br>(7 - 12) | 31<br>(26 - 35) | 1<br>(1 - 1) | 7<br>(-10 - 19) | 15<br>(12 - 18) | 4<br>(1 - 8) | 22<br>(18 - 25) |
| **≥80 years old** |  |  |  |  |  |  |  |  |  |
| Events observed included in impact estimation | 2023-2024 | 34,698 | 1,022 | 1,576 | 21,060 | 227 | 3,523 | 2,185 | 5,105 |
|  | 2024-2025 | 9,909 | 557 | 506 | 4,590 | 18 | 1,756 | 436 | 2,046 |
| NAE | 2023-2024 | 3,629<br>(3,089 - 4,432) | 149<br>(118 - 193) | 720<br>(619 - 861) | 526<br>(453 - 625) | 64<br>(51 - 85) | 698<br>(602 - 824) | 353<br>(268 - 470) | 1,119<br>(942 - 1,393) |
|  | 2024-2025 | 1,315<br>(1,008 - 1,621) | 55<br>(38 - 71) | 192<br>(147 - 238) | 61<br>(46 - 72) | 1<br>(-2 - 3) | 272<br>(208 - 331) | 20<br>(7 - 34) | 716<br>(546 - 885) |
| NAE / pop. size (per 100,000) | 2023-2024 | 58<br>(49 - 70) | 30<br>(24 - 39) | 275<br>(237 - 329) | 13<br>(11 - 15) | 186<br>(149 - 246) | 309<br>(267 - 365) | 53<br>(40 - 71) | 211<br>(177 - 262) |
|  | 2024-2025 | 20<br>(15 - 25) | 9<br>(6 - 12) | 69<br>(53 - 86) | 1<br>(1 - 2) | 2<br>(-5 - 9) | 105<br>(80 - 128) | 3<br>(1 - 5) | 122<br>(93 - 151) |
| PF (%) | 2023-2024 | 9<br>(8 - 11) | 13<br>(10 - 16) | 31<br>(28 - 35) | 2<br>(2 - 3) | 22<br>(18 - 27) | 17<br>(15 - 19) | 14<br>(11 - 18) | 18<br>(16 - 21) |
|  | 2024-2025 | 12<br>(9 - 14) | 9<br>(6 - 11) | 28<br>(22 - 32) | 1<br>(1 - 2) | 3<br>(-11 - 15) | 13<br>(11 - 16) | 4<br>(2 - 7) | 26<br>(21 - 30) |
| COVID-19-related death |  |  |  |  |  |  |  |  |  |
| **≥80 years old** |  |  |  |  |  |  |  |  |  |
| Events observed<br>included in impact<br>estimation | 2023-2024 | 5,604 | .. | 302 | 2,793 | 11 | 455 | 1,473 | 570 |
|  | 2024-2025 | 1,190 | .. | 51 | 517 | 0 | 40 | 428 | 154 |
| NAE | 2023-2024 | 811<br>(559 - 1,028) | .. | 188<br>(132 - 241) | 69<br>(45 - 85) | 7<br>(3 - 11) | 144<br>(104 - 175) | 208<br>(117 - 293) | 191<br>(134 - 246) |
|  | 2024-2025 | 156<br>(98 - 208) | .. | 27<br>(14 - 39) | 8<br>(5 - 10) | 0<br>(0 - 0) | 6<br>(2 - 8) | 32<br>(6 - 65) | 81<br>(49 - 115) |
| NAE / pop. size<br>(per 100,000) | 2023-2024 | 13<br>(9 - 16) | .. | 72<br>(50 - 92) | 2<br>(1 - 2) | 21<br>(10 - 32) | 64<br>(46 - 78) | 31<br>(18 - 44) | 36<br>(25 - 46) |
|  | 2024-2025 | 2<br>(1 - 3) | .. | 10<br>(5 - 14) | 0<br>(0 - 0) | 0<br>(0 - 0) | 2<br>(1 - 3) | 5<br>(1 - 10) | 14<br>(8 - 20) |
| PF (%) | 2023-2024 | 13<br>(9 - 15) | .. | 38<br>(30 - 44) | 2<br>(2 - 3) | 40<br>(23 - 50) | 24<br>(19 - 28) | 12<br>(7 - 17) | 25<br>(19 - 30) |
|  | 2024-2025 | 12<br>(8 - 15) | .. | 34<br>(22 - 44) | 2<br>(1 - 2) | - | 13<br>(5 - 17) | 7<br>(1 - 13) | 34<br>(24 - 43) |

NAE: number of averted events; PAE: prevented fraction; Values are shown as median (95% prediction interval) across 1000 repetitions

In the 2024–2025 study period, the vaccination impact in the participating countries/regions was considerably smaller regarding numbers of hospitalisation averted, but consistent in terms of PF: 859 (686–1,033) hospitalisations were averted in individuals aged 65–79 years and 1,315 (1,008–1,621) in individuals aged ≥80 years (median PF were 11% and 12%). Higher rates of averted hospitalisations were observed in Norway (41 per 100,000 inhabitants in the 65–79 age group) and Sweden (122 per 100,000 inhabitants in the ≥80 years age group) and low or approaching zero rates of averted hospitalisations in Italy, Navarre, Belgium and Portugal (0–9 per 100,000 inhabitants) (Table 2).

The estimated number of COVID-19 related deaths averted by vaccination was only possible to obtain for the population aged ≥80 years old. In this age group, and across all study sites, 811 (559–1,028) deaths were averted during the 2023–2024 study period and 156 (98–208) deaths were averted during the 2024–2025 study period (median PF were 13% and 12%). The rate of averted events per 100,000 individuals, varied from 2 in Italy to 72 in Denmark in the 2023-2024 study period and from close to zero in Italy and Navarre to 14 per 100,000 inhabitants in Sweden in the 2024-2025 study period (Table 2).

For the 2023–2024 study period, across all sites and both age groups, weekly rates of averted events peaked in December 2023–January 2024, then gradually declined despite the rise of the COVID-19 hospitalisations and deaths in the summer 2024. For the 2024–2025 study period, the highest weekly rate of events averted by vaccination peaked in December 2024–January 2025, then gradually declined to residual or null values during the spring and summer 2025. During both study periods, the highest weekly estimates were observed in Nordic countries (Figure S7 to S12).

Sensitivity analyses using 8-week pooled VE estimates (without interpolation) yielded results similar to those based on interpolated weekly VE, with a slightly lower vaccination impact across most study sites (Table S7 and S8).

## Discussion

Our study quantifies the public health impact of the 2023 and 2024 autumn COVID-19 vaccination campaigns in older adults across several EU/EEA countries. Overall, among adults aged ≥65 years, the 2023 autumn vaccination campaign prevented 6,200 COVID-19 hospitalisations vs 2,200 hospitalisations for the 2024 autumn vaccination campaign. For those aged ≥80 years old, the number of averted COVID-19 related deaths by the two vaccination campaigns were 811 and 156, respectively.

We observed a higher impact, in terms of numbers and rates of averted events, of the 2023 than the 2024 autumn COVID-19 vaccination campaigns in VEBIS-EHR countries/regions, although the prevented fractions were similar (9–13% in 2023 vs 11–12% in 2024).

Three main factors explain the lower impact of the 2024 autumn campaign. First, vaccine coverage was generally lower than in 2023. Second, COVID-19 hospitalisation and mortality rates were lower in the 2024–2025 study period. Third, the timing of peak incidence was suboptimal in 2024–2025: the highest rates occurred either before vaccine coverage reached its maximum or later, or when VE had waned (spring/summer). Having high vaccine coverage and high VE at times of high incidence helps maximise vaccination campaign impact, which was not the case for the 2024–2025 study period, compared with the 2023-2024 study period.

We also observed differences in the impact of the autumn vaccination campaigns across VEBIS-EHR countries/regions. The main drivers of these differences were the differences in vaccine coverage (10–85% for the 2023–2024 study period and 4–83% for the 2024–2025 study period) and the timing of vaccination roll-out and COVID-19 rates observed between study sites. Countries with high vaccine coverage and high VE during periods of high incidence (e.g. Denmark, Norway in the 2023–2024 study period) experienced the largest reductions in severe outcomes. Lower impact was observed in countries with low vaccine coverage (Italy) or with low COVID-19 hospitalisation or death rates (Portugal and Belgium). Sub-optimal impact was also identified when vaccine coverage peaked after epidemic decline (e.g. Sweden, Norway in the 2024–2025 study period) or when the peak of infections occurred after VE had already waned (summer 2024 and 2025).

To our knowledge, this is the first multi-country assessment of COVID-19 vaccination impact across Europe, and across several years, since the analyses covering the 2020–2021 period early in the pandemic (8). Compared with these previous studies, the impact was smaller, reflecting lower incidence, and lower VE and vaccine coverage than the levels observed during the early phases of the primary COVID-19 vaccination and following first and second booster doses (8).

A distinction between this study and those conducted during earlier pandemic phases is also the absence of widespread non-pharmaceutical interventions (NPI). During earlier phases, public health restrictions may have reduced disease transmission independently of vaccination (23). In addition, changes in SARS-CoV-2 testing strategies over time, the circulation of successive variants with differing transmissibility and virulence (24,25), and the accumulation of population immunity from both infection and vaccination have shaped observed trends in hospitalisations, mortality and VE. Our findings therefore offer a clearer view of the direct contribution of vaccination in a post-pandemic context with minimal population-level interventions and underlying population immunity.

This study has limitations. The use of routinely collected data may introduce bias in VE and severe outcome estimates. Unmeasured confounding could overestimate or underestimate impact, while underreporting of severe events in some sites may underestimate it (26). Site-specific data issues (e.g. coding delays in Portugal (26), or hospital admission data specificities in Belgium – see supplementary material – section 2) may have affected incidence estimates. However, as these limitations likely affected both vaccination campaigns similarly, observed differences between study periods are likely robust. The methods we applied did not account for indirect vaccination effect, so our results must be interpreted as a lower bound of the overall impact of vaccination. Nevertheless, we can assume that given the low vaccine coverage in other age groups, the indirect effect is likely to be low (27,28).

The present study showed the critical importance of vaccination timing with the expected peak of the COVID-19 incidence. The maximum impact of the vaccination campaign is achieved when the maximum vaccine coverage occurs during the period of highest COVID-19 incidence and at the VE maximum level, before it starts to decline. This situation was achieved in the majority of the VEBIS-EHR study sites during the first months of the 2023 autumn vaccination campaign, but it was not achieved during the first months of the 2024 autumn vaccination campaign, neither during the summer surges of COVID-19 in 2024 and 2025.

### Public health implications and conclusions

COVID-19 vaccination among adults ≥65 years continues to prevent substantial numbers of severe outcomes, though impact varies between years and countries/regions. Aligning vaccination timing with anticipated increases in viral circulation is essential to maximise impact, but there are challenges to do so, as seasonality is not yet established. Recent COVID-19 temporal patterns show summer waves and low incidence during autumn and winter (occurring in 2024/2025 and currently being observed again for 2025/2026), but it is unclear whether this pattern will continue.

Although coordination with seasonal influenza vaccination strategies is critical, given overlapping target populations, programmatic ease (i.e. timing of these two vaccination programmes) and to reduce winter pressure on health services, the current reduced autumn and winter incidence of COVID-19 reduces the impact of autumn COVID-19 vaccination programmes. Given that studies conducted in countries that implement spring vaccination campaigns (29) show that COVID-19 VE is restored to the observed levels of autumn/winter protection, and if the temporal pattern of COVID-19 incidence observed since the spring/summer of 2024 is maintained, a review of the current COVID-19 vaccination programme strategies should be discussed, in order to maximise the impact and cost-effectiveness of the current COVID-19 vaccination programmes and strategies.

Given the still uncertain temporal patters of COVID-19, ongoing monitoring of VE and vaccination programme impact through description of the temporal distribution of COVID-19 incidence, vaccine coverage and VE remains necessary to optimise vaccination timing, improve communication strategies and inform modelling, cost-effectiveness analyses, and resource allocation.

## Data Availability

Authors cannot share the data used for this study, which should be requested to the data owner institutions following their respective procedures.

