## Supplementary Material for "Impact of autumn 2023 and 2024 COVID-19 vaccination in preventing COVID-19 related hospitalisations and deaths in seven EU/EEA countries: a VEBIS-EHR network study"

Tables

Figures

### 1 - Ethics

**Ethics**

All study sites participating in this study conformed with their respective national and EU ethical and data protection requirements. Ethical statements for each of the participating study sites:

**Belgium:** Data linkage and collection within the data-warehouse have been approved by the information security committee. The study was conducted in accordance with the Declaration of Helsinki. Ethical approval was granted for the gathering of data from hospitalized patients by the Committee for Medical Ethics from the Ghent University Hospital (reference number BC-07507) and authorization for possible individual data linkage using the national register number from the Information Security Committee (ISC) Social Security and Health (reference number IVC/KSZG/20/384). Linkage of hospitalized patient data to vaccination and testing within the LINK-VACC project was approved by the Medical Ethics Committee UZ Brussels–VUB on 3 February 2021 (reference number 2020/523), and authorization from the ISC Social Security and Health (reference number IVC/KSZG/21/034).

**Denmark:** Only administrative register data was used for the study. According to Danish law, ethics approval is exempt for such research, and the Danish Data Protection Agency, which is dedicated ethics and legal oversight body, thus waives ethical approval for the study of administrative register data when no individual contact of participants is necessary, and only aggregate results are included as findings. The study is, therefore, fully compliant with all legal and ethical requirements, and there are no further processes available regarding such studies.

**Navarre (Spain):** The study was approved by Navarre’s Ethical Committee for Clinical Research (PI_2024/150), which waived the requirement of obtaining informed consent.

**Norway:** The Norwegian Institute of Public Health (NIPH) has statutory responsibilities both as a national producer of scientific knowledge and as the authority responsible for managing the mandated health registries. For defined purposes, selected subsets of data from these registries are integrated—made accessible through the Stat19 framework—to generate population-level insights and evidence in the form of anonymous statistics. According to Section 19 of the Health Registry Act, FHI is explicitly authorised and obligated to combine registry data for the production of relevant statistics.

**Portugal:** The study received approval from the Ethical Committee and the Data Protection Officer of the Instituto Nacional de Saúde Doutor Ricardo Jorge. Given that data was irreversibly anonymised, the need for the participants’ informed consent was waived by the Ethical Committee.

**Italy:** This study, based on routinely collected data, will not be submitted for approval to an ethical committee because the dissemination of COVID-19 surveillance data was authorized by the Italian law N. 52 of 19 May 2022, following the law decree N. 24 of 24 March 2022 (Article n. 13). Based on the same acts, the information on COVID-19 vaccination was retrieved by the Italian National Institute of Health using data from the National Immunisation Information System of the Italian Ministry of Health. Because of the retrospective design and the large size of the population under study, in accordance with the Authorization n. 9 released by the Italian data protection authority on 15 December 2016, the individual informed consent was not requested for the conduction of this study.

**Sweden:** The Swedish study is approved by the Swedish Ethical Review Authority (2020-06859, 2021-02186) and has conformed to the principles embodied in the Declaration of Helsinki. Consent to participate is not applicable as this is a register-based study.

### 2 - Specificities of Belgium hospitalisation data and interpretation of their estimates

The weekly number of COVID-19 hospitalisations in the eligible population of the study cohort as reported by study sites was used, with the exception of Belgium.

Belgium uses the CHS (Clinical Hospital Surveillance) dataset for VE studies within VEBIS-EHR. However, CHS underreports COVID-19 hospitalisations, leading to underestimated rates. Sciensano also manages the SARI (Severe Acute Respiratory Infections) database, which is exhaustive for participating hospitals and can be used to estimate total hospitalisations nationwide. Since SARI is not integrated into the LINK-VACC environment, it cannot directly be used for VE calculations as defined in the VEBIS protocol (more information on LINK-VACC is available here: https://www.sciensano.be/en/projects/linking-registers-covid-19-vaccine-surveillance).

As a consequence, VE was estimated using CHS, while SARI was used to estimate the incidence of COVID-19 hospitalisations in Belgium. The SARI network, originally designed for influenza, was expanded on 13 November 2023 to 10 hospitals (9 since November 2024), with its case definition adapted to capture other respiratory infections, including COVID-19.

For each hospitalisation, SARI collects demographic and clinical information, risk factors, vaccination status, and virological results from respiratory samples tested for 16 viruses (including SARS-CoV-2, influenza, and RSV). While surveillance was seasonal before 2020 and incomplete until late 2023, the digitalisation of the system through a RedCap questionnaire on 13 November 2023 enabled continuous reporting and full coverage from early 2024 onwards.

National estimates of COVID-19 hospitalisations are derived by adjusting the number of SARS-CoV-2 positive SARI hospitalisations to the Belgian population, using the catchment population of the participating hospitals, based on municipality-level hospital market shares.

**Interpretation of Belgium results**

Several factors should be considered when interpreting the Belgian results.
Firstly, to match VEBIS-EHR selection criteria, nursing home residents were excluded from the numerator (cases). But they could not be excluded from the denominator (catchment population), due to the lack of up-to-date information allowing for their exclusion. This asymmetry may have led to an underestimation of the hospitalisation rate.
Secondly, the SARI case definition applied for this analysis was a stricter version, which required both fever (≥38°C) and respiratory symptoms (cough and/or shortness of breath) of sudden onset within 10 days prior to admission. This definition was replaced on 12 November 2023 with a broader one, more appropriate to the COVID-19 clinical presentation, as fever is not consistently present in COVID-19 cases. The use of the older definition may therefore have underestimated the number of COVID-19 hospitalisations captured by SARI.
Finally, the SARI network functions as a sentinel surveillance system. Before its expansion in late 2023, the number of reporting hospitals was limited, and the system’s sensitivity may have been lower. Although coverage improved substantially after November 2023, estimates prior to this period should be interpreted with caution.

### 3 - Interpolating weekly VE from 8-week VE

We used 8-week VE estimates to estimate VE at the weekly level using an interpolation method. The 8-week VE was modelled by either an exponential or a logistic decay function, allowing for a slower early decay and more flexibility overall, depending on which one provides the best fit as assessed by the Bayesian information criterion (BIC). The model fit was assessed on the (log(1-VE)) (or log aHR) scale because it is the scale at which the estimates and their standard errors are primarily calculated (Figure below).

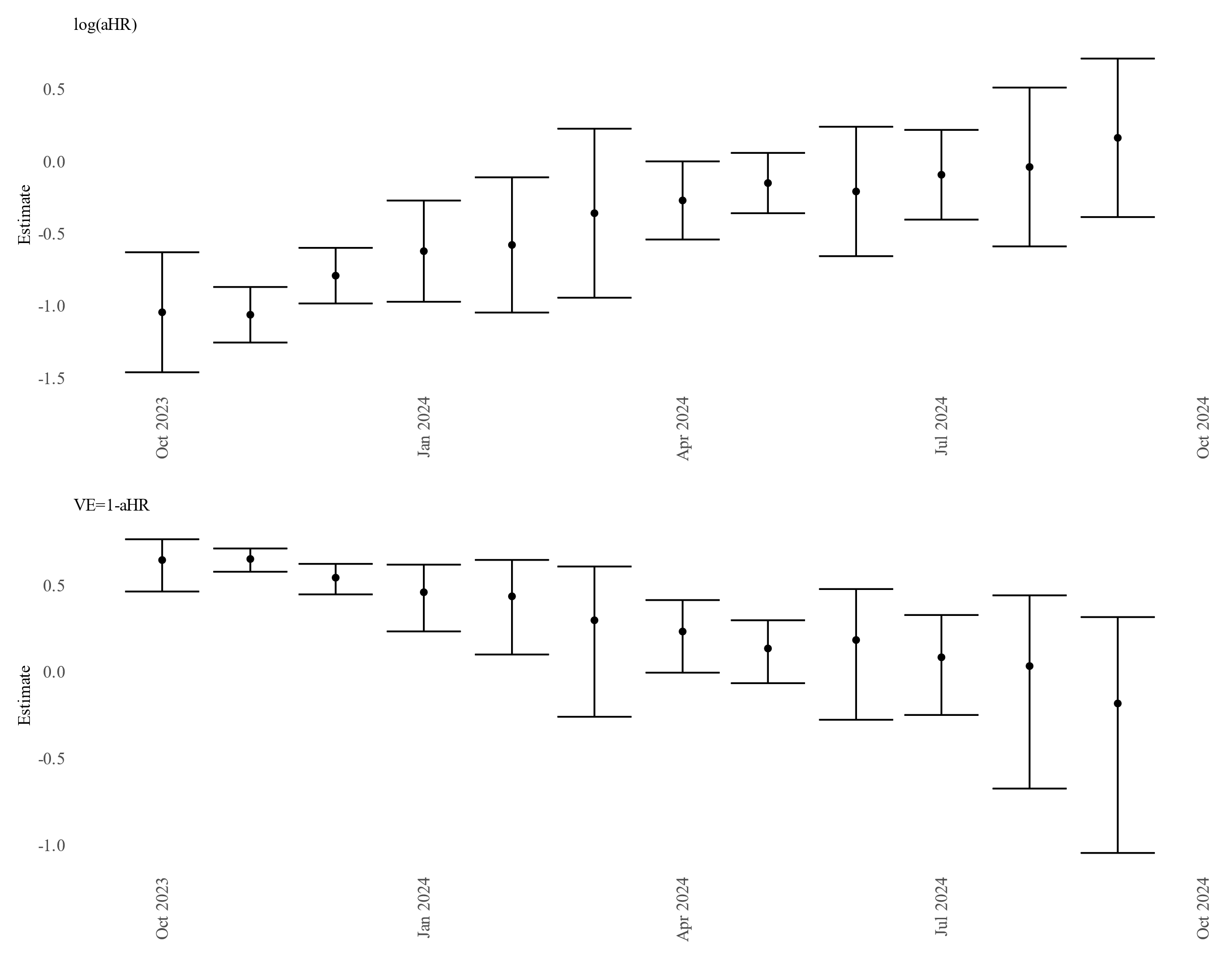

Figure S1: Illustration of vaccine effectiveness estimates (VE), their 95% confidence intervals (the error bars), and their corresponding values on the original scale of log(aHR) using mock estimates.

An exponential decay function is as follows: ${VE}_{t}={VE}_{t=0}\times e^{-b\times t}$ with *t* an indicator for the week and $b>0$ is the decay rate. A logistic decay function is as follows: ${VE}_{t}=\frac{{VE}_{t=0}}{1+a\times e^{b\times t}}$ with *t* an indicator for the week, $a$ is constant related with the initial conditions and $b>0$ is the decay rate.

We fitted both decay functions to our estimates produced monthly over rolling 8-week periods on the log(aHR) scale through non-linear regression technique, by minimising the sum of squared residuals.

The decay functions rearranged on the log(aHR) before applying non-linear regression techniques are as follows:

Exponential decay ${log(aHR}_{t})=log(1-{VE}_{t=0}\times e^{-b\times t})$

Logistic decay: ${log(aHR}_{t})=log(1-\frac{{VE}_{t=0}}{1+a\times e^{b\times t}})$

The model parameters were estimated by minimising, for each 8-week period, the difference between the observed 8-week VE and the weekly VE estimation at the middle of the corresponding 8-week period (in the log HR scale).

The predicted values on the log(aHR) scale were then transformed to the VE scale to provide weekly estimates.

**Resampling procedure**

To account for the uncertainty around the VE estimates, we implemented a resampling procedure (1,000 repetitions).

For each iteration r, the following steps were performed:

1. A random sample of 8-week VE values was generated based on the observed VE estimates and their corresponding standard errors.
2. A logistic or exponential decay model was fitted to these sampled data; the estimated decay parameters and the residual standard error SEr were stored for iteration r.
3. Using the parameters obtained in step 2, a trajectory of weekly VE was predicted for the full study period.
4. Additional uncertainty was incorporated at the weekly level according to:

logHR_with_uncertainty ~ N(logHR, SE_r_)

5. The impact indicators were then computed using the weekly VE estimates obtained in step 4.

For the sensitivity analysis, we used the samples generated at step 1.

### Tables

Table S1: 2023 and 2024 autumn COVID-19 vaccination campaigns dates, VEBIS-EHR, Europe

| Belgium | 11^th^ September 2023 | 23^rd^ September 2024 |
| --- | --- | --- |
| Denmark | 1^st^ October 2023 | 1^st^ October 2024 |
| Italy | 1^st^ October 2023 | 18^th^ September 2024 |
| Sweden | 1^st^ September 2023 | 15^th^ October 2024 |
| Navarre (Spain) | 16^th^ October 2023 | 14^th^ October 2024 |
| Norway | 1^st^ October 2023 | 30^th^ September 2024 |
| Portugal | 29^th^ September 2023 | 20^th^ September 2024 |

Table S2: Study site specific criteria used to define low, medium, and high-risk comorbidities

|  | **Belgium** | | **Denmark** | **Italy** | **Navarre (Spain)** | **Norway** | **Portugal** | **Sweden** |
| --- | --- | --- | --- | --- | --- | --- | --- | --- |
| **Low risk** |  | No comorbidities associated with an increased risk for severe COVID-19 infection | | | | | | |
| **Medium risk**  **or**  **High risk** | At least one comorbidity which increases the risk for severe COVID-19 infection and not being immunocompromised (medium risk):   - *Received multidisciplinary oncologic consult* - *Cardiovascular illness – general* - *Cardiovascular illness- specifically a heart disease* - *Alzheimer* - *Asthma* - *Haemophilia* - *Chronic obstructive pulmonary disease* - *Diabetes with cardiovascular compilations* - *Diabetes Mellitus with insulin treatment* - *Epilepsy and neuropathic pain* - *Chronic hepatitis type B or C* - *Kidney failure* - *Cystic fibrosis* - *Exocrine pancreatic disease* - *Disease of Parkinson* - *Psychosis occurring with people older than 70 years* - *Psychosis occurring with people of 70 year or younger.* - *Multiple sclerosis* - *Thrombosis while treated with antithrombotic medicines* - *Thyroid disorder* - *HIV*   Immunocompromised (high risk): *if a person received a priority invitation for a COVID-19 vaccination due to being immunocompromised, then he/she was classified into this group but* he/she does not belong to our list of immunocompromising conditions used to identify ICPs | | Other, including:   - *Diabetes* - *Obesity* - *Neurological Disease* - *Kidney disease* - *Heart disease* - *Chronic respiratory disease* - *Liver disease (incl. alcohol lever)* - *Endocrine Disease* - *Hematological Disease* - *Coagulation Disease* - *Innate Diseases* - *TB* - *Missing a lung* - *Missing a kidney*   Immunocompromised, but does not belong to our list of immunocompromising conditions used to identify ICPs, including:   - *HIV* - *Immunological disease* | Other comorbidities, including:   - *Respiratory diseases requiring oxygen therapy, idiopathic pulmonary fibrosis* - *Advanced heart failure (Classes III-IV NYHA) and post cardiogenic shock patients* - *Amyotrophic lateral sclerosis and other motor neuron disorders, multiple sclerosis, muscular dystrophy, infantile cerebral palsy, myasthenia gravis, dysimmune neuropathies* - *Type 1 diabetes, Type 2 diabetes with complications or requiring combination therapy (with at least two anti-diabetes drugs)* - *Addison's disease* - *Panhypopituitarism* - *Cystic fibrosis* - *Intracerebral ischemic or hemorrhagic event that has led to impaired neurological and cognitive autonomy* - *Individuals who have had a stroke on 2020 or later ranked as level 3 or higher* - *Thalassemia major* - *Sickle cell anemia* - *Other severe anemias* - *Down syndrome* - *Body Mass Index >35* - *Severely disabled persons pursuant to law 104/1992 art. 3 paragraph 3* - *Chronic Alcohol Misuse* - *Functional or anatomic asplenia* - *COPD* - *Coagulopathies* - *Diabetes Mellitus and other endocrinopathies* - *Patients in hemodialysis or with chronic kidney diseases expected to start dialysis* - *Hemoglobinopathy such as sickle cell anemia or thalassemia* - *Chronic Liver Disease* - *Cochlear implant* - *Chronic Kidney Disease* - *Chronic eczema* - *Diseases associated with a high risk of aspiration pneumonia* - *Chronic Cardiovascular Disease* - *Chronic Respiratory Disease* - *Motor neuron diseases* - *Chronic inflammatory diseases and malabsorption syndromes* - *Obesity (Body Mass Index 30-35)* - *Drug Misuse* - *Patients with CSF leak from trauma or intervention* - *Metabolic diseases* - *Hematopoietic diseases* - *Pathologies that require important surgical interventions* - *Neurological diseases* - *Cerebrovascular diseases* - *Down Syndrome*   *Disabilities (physical, sensorial, learning or psychic)*  Immunocompromised but does not belong to our list of immunocompromising conditions, including:   - *Immunocompromised defects of the complement system* - *Other specified disorders involving the immune mechanism* - *Deficiency or dysfunction of a single component (C1-C9)* - *Deficiency of cell-mediated immunity* - *Deficiency of humoral immunity* - *Human immunodeficiency virus [HIV] disease, Human immunodeficiency virus, type 2 [HIV-2], Asymptomatic human immunodeficiency virus [HIV] infection status* - *Disorders involving the immune mechanism* - *Congenital and acquired disorders with poor antibody production* | Other major chronic conditions   - *Diabetes* - *Severe Obesity* - *Ictus* - *Dementia* - *Kidney disease* - *Heart disease* - *Chronic respiratory disease* - *Liver disease*   Immunocompromised but does not belong to our list of immunocompromising conditions used to identify ICPs | Medium risk:  - Chronic liver disease or significant hepatic impairment  -Diseases requiring immunosuppressive therapy  -Diabetes  -Chronic lung disease including cystic fibrosis and severe asthma which have required the use of high dose inhaled or oral steroids within the past year  - Obesity with a body mass index (BMI) of ≥35 kg/m2  - Dementia  - Chronic heart and vascular disease (with the exception of high blood pressure) and stroke  High risk:  - Organ transplant  -Immunodeficiency  -Haematological cancer in the last five years  -Other active cancers  -Neurological or neuromuscular diseases that cause impaired cough or lung function (e.g., ALS and cerebral palsy)  -Chronic kidney disease, or significant renal impairment. | Comorbidities that do not belong to our list of immunocompromising conditions used to identify ICPs  *Considered comorbidities include: anemia, asthma, cardiac disease, dementia, diabetes, hypertension, HIV, liver disease, neuromuscular disease, obesity, pulmonary disease, renal disease, rheumatologic disease, stroke, tuberculosis* | - Vaccine priority groups: LISA Healthcare worker (status per October 2018), SOL Nursing home resident (status per December 31, 2020). - Comorbidity groups: (binary) Any records with ICD-10 codes as primary/secondary diagnosis from inpatient stay or outpatient contact in hospital or from private practicing specialists, January 1, 2017 – December 27, 2020) - Chronic pulmonary disease NPR J41 J42 J43 J44 J45 J46 J47 J84 J98 E84 - Cardiovascular conditions and diabetes NPR, SPDR I05 I06 I07 I08 I09 I110 I2 I34 I35 I36 I37 I39 I42 I43 I46 I48 I49 I50 E10-E14 - ATC: A10 (at least two filled prescriptions during 2020, before December 27, 2020) - Autoimmunity-related conditions NPR D86 G35 K50 K51 L40 M05 M06 M07 M08 M09 M13 M14 M45 - Moderate to severe renal disease NPR I12 I13 N00 N01 N02 N03 N04 N05 N07 N11 N14 N17 N18 N19 Q61 |

Table S3: Definition of the 8-week periods for which VE against hospitalisation/death was estimated at study site level and the corresponding weeks covered by the VE estimates.

| **2023-2024 autumn vaccination campaign** | | **2024-2025 autumn vaccination campaign** | |
| --- | --- | --- | --- |
| **Eight-week follow up period** | **Weeks covered** | **Eight-week follow up period** | **Weeks covered** |
| October 1 to November 25, 2023 | 2023-W40 to 2023-W47 | October 1 to November 25, 2024 | 2024-W40 to 2024-W48 |
| November 1 to December 26, 2023 | 2023-W44 to 2023-W51 | November 1 to December 26, 2024 | 2024-W44 to 2024-W52 |
| December 1, 2023, to January 25, 2024 | 2023-W49 to 2024-W04 | December 1, 2024, to January 25, 2025 | 2024-W48 to 2025-W04 |
| January 1 to February 25, 2024 | 2024-W01 to 2024-W08 | January 1 to February 25, 2025 | 2025-W01 to 2025-W09 |
| February 1 to March 27, 2024 | 2024-W05 to 2024-W13 | February 1 to March 27, 2025 | 2025-W05 to 2025-W13 |
| March 1 to April 25, 2024 | 2024-W09 to 2024-W17 | March 1 to April 25, 2025 | 2025-W09 to 2025-W17 |
| April 1 to May 26, 2024 | 2024-W14 to 2024-W22 | April 1 to May 26, 2025 | 2025-W14 to 2025-W22 |
| May 1 to June 25, 2024 | 2024-W18 to 2024-W26 | May 1 to June 25, 2025 | 2025-W18 to 2025-W26 |
| June 1 to July 26, 2024 | 2024-W22 to 2024-W30 | June 1 to July 26, 2025 | 2025-W22 to 2025-W30 |
| July 1 to August 25, 2024 | 2024-W27 to 2024-W35 | July 1 to August 25, 2025 | 2025-W27 to 2025-W35 |
| August 1 to September 25, 2024 | 2024-W31 to 2024-W39 | August 1 to September 25, 2025 | 2025-W31 to 2025-W39 |
| September 1 to October 26, 2024 | 2024-W36 to 2024-W43 | September 1 to October 26, 2025 | 2025-W36 to 2025-W43 |

Table S4: Interpolation BIC estimates

| **Study period** | **Outcome** | **Age group** | **BIC exponential model** | **BIC logistic model** |
| --- | --- | --- | --- | --- |
| **2023-2024** | **Hospitalisation** | 65–79 years | -15.41 | -33.22 |
|  |  | ≥80 years | -11.98 | -14.23 |
|  | **Deaths** | 65–79 years | Not estimated due to missing values in 8-week VE data | Not estimated due to missing values in 8-week VE data |
|  |  | ≥80 years | 3.25 | -16.01 |

| **Study period** | **Outcome** | **Age group** | **BIC exponential model** | **BIC logistic model** |
| --- | --- | --- | --- | --- |
| **2024-2025** | **Hospitalisation** | 65–79 years | -1.77 | -11.46 |
|  |  | ≥80 years | -12.29 | -19.07 |
|  | **Deaths** | 65–79 years | Not estimated due to missing values in 8-week VE data | Not estimated due to missing values in 8-week VE data |
|  |  | ≥80 years | -2.69 | -4.25 |

Table S5: Weekly vaccine effectiveness estimates from the logistic decay interpolation model of the eight-week vaccine effectiveness estimates, 2023–2024, VEBIS-EHR, Europe.

| **Week** | **Interpolated Weekly VE (median – 95% prediction interval) – Hospitalisation due to COVID-19 – 65-79 years old** | **Interpolated Weekly VE (median – 95% prediction interval) – Hospitalisation due to COVID-19 – ≥80 years old** | **Interpolated Weekly VE (median – 95% prediction interval) –COVID-19-related deaths – ≥80 years old** |
| --- | --- | --- | --- |
| 2023-W40 | 0.66 (0.53 - 0.77) | 0.67 (0.48 - 0.78) | 0.60 (0.03 - 0.83) |
| 2023-W41 | 0.66 (0.54 - 0.76) | 0.66 (0.49 - 0.76) | 0.60 (0.21 - 0.82) |
| 2023-W42 | 0.65 (0.52 - 0.75) | 0.65 (0.48 - 0.75) | 0.60 (0.16 - 0.80) |
| 2023-W43 | 0.65 (0.50 - 0.75) | 0.64 (0.47 - 0.74) | 0.60 (0.18 - 0.81) |
| 2023-W44 | 0.64 (0.51 - 0.74) | 0.63 (0.47 - 0.73) | 0.59 (0.11 - 0.80) |
| 2023-W45 | 0.63 (0.52 - 0.73) | 0.62 (0.47 - 0.72) | 0.59 (0.14 - 0.80) |
| 2023-W46 | 0.62 (0.51 - 0.71) | 0.60 (0.46 - 0.71) | 0.59 (0.16 - 0.80) |
| 2023-W47 | 0.61 (0.48 - 0.70) | 0.59 (0.44 - 0.68) | 0.58 (0.04 - 0.80) |
| 2023-W48 | 0.60 (0.49 - 0.69) | 0.58 (0.43 - 0.68) | 0.58 (0.17 - 0.79) |
| 2023-W49 | 0.59 (0.45 - 0.68) | 0.56 (0.41 - 0.68) | 0.57 (0.14 - 0.80) |
| 2023-W50 | 0.57 (0.45 - 0.67) | 0.54 (0.41 - 0.66) | 0.56 (0.12 - 0.78) |
| 2023-W51 | 0.56 (0.42 - 0.66) | 0.53 (0.38 - 0.66) | 0.55 (0.05 - 0.78) |
| 2023-W52 | 0.54 (0.41 - 0.65) | 0.51 (0.37 - 0.64) | 0.55 (0.16 - 0.78) |
| 2024-W01 | 0.53 (0.38 - 0.64) | 0.49 (0.34 - 0.64) | 0.54 (0.03 - 0.78) |
| 2024-W02 | 0.51 (0.36 - 0.63) | 0.47 (0.32 - 0.64) | 0.54 (-0.02 - 0.77) |
| 2024-W03 | 0.50 (0.33 - 0.62) | 0.45 (0.28 - 0.61) | 0.52 (-0.04 - 0.77) |
| 2024-W04 | 0.47 (0.31 - 0.60) | 0.43 (0.27 - 0.60) | 0.52 (0.00 - 0.76) |
| 2024-W05 | 0.46 (0.28 - 0.60) | 0.41 (0.24 - 0.60) | 0.51 (0.06 - 0.75) |
| 2024-W06 | 0.43 (0.24 - 0.57) | 0.39 (0.22 - 0.59) | 0.48 (-0.05 - 0.76) |
| 2024-W07 | 0.41 (0.22 - 0.56) | 0.38 (0.19 - 0.57) | 0.49 (-0.02 - 0.75) |
| 2024-W08 | 0.40 (0.18 - 0.55) | 0.36 (0.16 - 0.55) | 0.48 (-0.12 - 0.74) |
| 2024-W09 | 0.37 (0.13 - 0.55) | 0.33 (0.13 - 0.56) | 0.45 (-0.14 - 0.74) |
| 2024-W10 | 0.35 (0.08 - 0.52) | 0.31 (0.08 - 0.54) | 0.44 (-0.19 - 0.74) |
| 2024-W11 | 0.33 (0.08 - 0.50) | 0.29 (0.09 - 0.50) | 0.42 (-0.12 - 0.72) |
| 2024-W12 | 0.31 (0.06 - 0.48) | 0.28 (0.03 - 0.48) | 0.43 (-0.19 - 0.71) |
| 2024-W13 | 0.29 (0.03 - 0.47) | 0.25 (0.01 - 0.49) | 0.40 (-0.31 - 0.72) |
| 2024-W14 | 0.26 (-0.02 - 0.45) | 0.24 (-0.02 - 0.45) | 0.37 (-0.32 - 0.69) |
| 2024-W15 | 0.25 (-0.03 - 0.44) | 0.22 (-0.01 - 0.46) | 0.36 (-0.27 - 0.68) |
| 2024-W16 | 0.22 (-0.06 - 0.44) | 0.20 (-0.08 - 0.44) | 0.35 (-0.40 - 0.69) |
| 2024-W17 | 0.21 (-0.09 - 0.42) | 0.19 (-0.04 - 0.43) | 0.34 (-0.43 - 0.68) |
| 2024-W18 | 0.20 (-0.14 - 0.40) | 0.17 (-0.10 - 0.41) | 0.33 (-0.43 - 0.65) |
| 2024-W19 | 0.17 (-0.10 - 0.37) | 0.15 (-0.10 - 0.40) | 0.30 (-0.55 - 0.63) |
| 2024-W20 | 0.16 (-0.16 - 0.37) | 0.14 (-0.16 - 0.39) | 0.27 (-0.54 - 0.64) |
| 2024-W21 | 0.15 (-0.12 - 0.36) | 0.13 (-0.13 - 0.36) | 0.28 (-0.58 - 0.64) |
| 2024-W22 | 0.13 (-0.15 - 0.34) | 0.12 (-0.14 - 0.37) | 0.26 (-0.52 - 0.63) |
| 2024-W23 | 0.12 (-0.16 - 0.37) | 0.11 (-0.19 - 0.36) | 0.24 (-0.53 - 0.66) |
| 2024-W24 | 0.10 (-0.14 - 0.30) | 0.09 (-0.19 - 0.32) | 0.20 (-0.58 - 0.60) |
| 2024-W25 | 0.09 (-0.18 - 0.32) | 0.09 (-0.21 - 0.33) | 0.20 (-0.65 - 0.60) |
| 2024-W26 | 0.09 (-0.19 - 0.33) | 0.08 (-0.24 - 0.34) | 0.20 (-0.69 - 0.62) |
| 2024-W27 | 0.08 (-0.18 - 0.30) | 0.08 (-0.21 - 0.33) | 0.17 (-0.68 - 0.61) |
| 2024-W28 | 0.08 (-0.21 - 0.30) | 0.07 (-0.21 - 0.32) | 0.18 (-0.84 - 0.58) |
| 2024-W29 | 0.07 (-0.22 - 0.29) | 0.06 (-0.26 - 0.30) | 0.17 (-0.71 - 0.60) |
| 2024-W30 | 0.06 (-0.18 - 0.28) | 0.05 (-0.22 - 0.31) | 0.15 (-0.76 - 0.58) |
| 2024-W31 | 0.05 (-0.22 - 0.26) | 0.05 (-0.26 - 0.31) | 0.14 (-0.83 - 0.58) |
| 2024-W32 | 0.05 (-0.23 - 0.25) | 0.04 (-0.23 - 0.28) | 0.12 (-0.73 - 0.56) |
| 2024-W33 | 0.05 (-0.20 - 0.29) | 0.05 (-0.23 - 0.28) | 0.13 (-0.80 - 0.58) |
| 2024-W34 | 0.04 (-0.28 - 0.26) | 0.04 (-0.28 - 0.27) | 0.11 (-0.83 - 0.56) |
| 2024-W35 | 0.04 (-0.26 - 0.25) | 0.04 (-0.29 - 0.28) | 0.12 (-0.82 - 0.57) |
| 2024-W36 | 0.04 (-0.24 - 0.28) | 0.03 (-0.30 - 0.31) | 0.12 (-0.80 - 0.59) |
| 2024-W37 | 0.04 (-0.25 - 0.25) | 0.03 (-0.28 - 0.26) | 0.11 (-0.83 - 0.56) |
| 2024-W38 | 0.03 (-0.26 - 0.24) | 0.03 (-0.29 - 0.25) | 0.09 (-0.91 - 0.57) |
| 2024-W39 | 0.03 (-0.25 - 0.24) | 0.03 (-0.30 - 0.27) | 0.09 (-0.85 - 0.57) |

Table S6: Weekly vaccine effectiveness estimates from the logistic decay interpolation model of the eight-week vaccine effectiveness estimates, 2024–2025, VEBIS-EHR, Europe.

| **Week** | **Interpolated Weekly VE (median – 95% prediction interval) – Hospitalisation due to COVID-19 – 65-79 years old** | **Interpolated Weekly VE (median – 95% prediction interval) – Hospitalisation due to COVID-19 – ≥80 years old** | **Interpolated Weekly VE (median – 95% prediction interval) –COVID-19-related deaths – ≥80 years old** |
| --- | --- | --- | --- |
| 2024-W40 | 0.61 (0.42 - 0.75) | 0.61 (0.40 - 0.74) | 0.60 (0.03 - 0.83) |
| 2024-W41 | 0.61 (0.42 - 0.74) | 0.60 (0.44 - 0.74) | 0.60 (0.21 - 0.82) |
| 2024-W42 | 0.61 (0.41 - 0.74) | 0.60 (0.39 - 0.72) | 0.60 (0.16 - 0.80) |
| 2024-W43 | 0.60 (0.42 - 0.74) | 0.59 (0.37 - 0.72) | 0.60 (0.18 - 0.81) |
| 2024-W44 | 0.60 (0.40 - 0.74) | 0.59 (0.37 - 0.71) | 0.59 (0.11 - 0.80) |
| 2024-W45 | 0.60 (0.38 - 0.73) | 0.58 (0.37 - 0.71) | 0.59 (0.14 - 0.80) |
| 2024-W46 | 0.60 (0.42 - 0.73) | 0.57 (0.40 - 0.70) | 0.59 (0.16 - 0.80) |
| 2024-W47 | 0.59 (0.38 - 0.73) | 0.56 (0.35 - 0.69) | 0.58 (0.04 - 0.80) |
| 2024-W48 | 0.59 (0.41 - 0.72) | 0.56 (0.37 - 0.69) | 0.58 (0.17 - 0.79) |
| 2024-W49 | 0.58 (0.39 - 0.73) | 0.54 (0.34 - 0.69) | 0.57 (0.14 - 0.80) |
| 2024-W50 | 0.58 (0.39 - 0.71) | 0.54 (0.35 - 0.67) | 0.56 (0.12 - 0.78) |
| 2024-W51 | 0.58 (0.35 - 0.71) | 0.53 (0.29 - 0.67) | 0.55 (0.05 - 0.78) |
| 2024-W52 | 0.57 (0.38 - 0.71) | 0.51 (0.31 - 0.66) | 0.55 (0.16 - 0.78) |
| 2025-W01 | 0.56 (0.34 - 0.71) | 0.50 (0.27 - 0.66) | 0.54 (0.03 - 0.78) |
| 2025-W02 | 0.56 (0.34 - 0.72) | 0.49 (0.26 - 0.65) | 0.54 (-0.02 - 0.77) |
| 2025-W03 | 0.54 (0.32 - 0.71) | 0.48 (0.23 - 0.64) | 0.52 (-0.04 - 0.77) |
| 2025-W04 | 0.54 (0.31 - 0.70) | 0.46 (0.23 - 0.62) | 0.52 (0.00 - 0.76) |
| 2025-W05 | 0.52 (0.30 - 0.68) | 0.45 (0.23 - 0.60) | 0.51 (0.06 - 0.75) |
| 2025-W06 | 0.51 (0.28 - 0.68) | 0.42 (0.19 - 0.59) | 0.48 (-0.05 - 0.76) |
| 2025-W07 | 0.50 (0.27 - 0.67) | 0.41 (0.17 - 0.58) | 0.49 (-0.02 - 0.75) |
| 2025-W08 | 0.49 (0.25 - 0.66) | 0.39 (0.13 - 0.57) | 0.48 (-0.12 - 0.74) |
| 2025-W09 | 0.47 (0.18 - 0.66) | 0.37 (0.07 - 0.56) | 0.45 (-0.14 - 0.74) |
| 2025-W10 | 0.46 (0.18 - 0.64) | 0.35 (0.06 - 0.54) | 0.44 (-0.19 - 0.74) |
| 2025-W11 | 0.44 (0.15 - 0.63) | 0.33 (0.05 - 0.53) | 0.42 (-0.12 - 0.72) |
| 2025-W12 | 0.43 (0.14 - 0.61) | 0.32 (0.03 - 0.51) | 0.43 (-0.19 - 0.71) |
| 2025-W13 | 0.41 (0.08 - 0.61) | 0.29 (0.00 - 0.49) | 0.40 (-0.31 - 0.72) |
| 2025-W14 | 0.38 (0.04 - 0.59) | 0.26 (-0.05 - 0.47) | 0.37 (-0.32 - 0.69) |
| 2025-W15 | 0.37 (0.07 - 0.59) | 0.25 (-0.06 - 0.47) | 0.36 (-0.27 - 0.68) |
| 2025-W16 | 0.34 (-0.02 - 0.57) | 0.23 (-0.14 - 0.45) | 0.35 (-0.40 - 0.69) |
| 2025-W17 | 0.33 (-0.02 - 0.56) | 0.21 (-0.14 - 0.46) | 0.34 (-0.43 - 0.68) |
| 2025-W18 | 0.31 (-0.07 - 0.54) | 0.19 (-0.15 - 0.42) | 0.33 (-0.43 - 0.65) |
| 2025-W19 | 0.29 (-0.09 - 0.52) | 0.18 (-0.16 - 0.42) | 0.30 (-0.55 - 0.63) |
| 2025-W20 | 0.25 (-0.14 - 0.50) | 0.15 (-0.21 - 0.41) | 0.27 (-0.54 - 0.64) |
| 2025-W21 | 0.25 (-0.14 - 0.49) | 0.15 (-0.21 - 0.39) | 0.28 (-0.58 - 0.64) |
| 2025-W22 | 0.24 (-0.15 - 0.50) | 0.14 (-0.19 - 0.38) | 0.26 (-0.52 - 0.63) |
| 2025-W23 | 0.20 (-0.15 - 0.49) | 0.11 (-0.24 - 0.39) | 0.24 (-0.53 - 0.66) |
| 2025-W24 | 0.18 (-0.19 - 0.45) | 0.10 (-0.25 - 0.34) | 0.20 (-0.58 - 0.60) |
| 2025-W25 | 0.17 (-0.27 - 0.46) | 0.09 (-0.29 - 0.35) | 0.20 (-0.65 - 0.60) |
| 2025-W26 | 0.17 (-0.27 - 0.45) | 0.09 (-0.28 - 0.36) | 0.20 (-0.69 - 0.62) |
| 2025-W27 | 0.15 (-0.24 - 0.44) | 0.07 (-0.26 - 0.35) | 0.17 (-0.68 - 0.61) |
| 2025-W28 | 0.13 (-0.32 - 0.41) | 0.07 (-0.34 - 0.33) | 0.18 (-0.84 - 0.58) |
| 2025-W29 | 0.12 (-0.30 - 0.42) | 0.07 (-0.37 - 0.34) | 0.17 (-0.71 - 0.60) |
| 2025-W30 | 0.11 (-0.34 - 0.38) | 0.06 (-0.34 - 0.32) | 0.15 (-0.76 - 0.58) |
| 2025-W31 | 0.09 (-0.36 - 0.40) | 0.05 (-0.32 - 0.31) | 0.14 (-0.83 - 0.58) |
| 2025-W32 | 0.08 (-0.35 - 0.38) | 0.04 (-0.31 - 0.30) | 0.12 (-0.73 - 0.56) |
| 2025-W33 | 0.08 (-0.33 - 0.38) | 0.05 (-0.33 - 0.31) | 0.13 (-0.80 - 0.58) |
| 2025-W34 | 0.07 (-0.41 - 0.38) | 0.04 (-0.40 - 0.31) | 0.11 (-0.83 - 0.56) |
| 2025-W35 | 0.07 (-0.37 - 0.36) | 0.04 (-0.31 - 0.31) | 0.12 (-0.82 - 0.57) |
| 2025-W36 | 0.06 (-0.42 - 0.37) | 0.04 (-0.34 - 0.31) | 0.12 (-0.80 - 0.59) |
| 2025-W37 | 0.05 (-0.42 - 0.38) | 0.03 (-0.34 - 0.32) | 0.11 (-0.83 - 0.56) |
| 2025-W38 | 0.04 (-0.40 - 0.35) | 0.03 (-0.37 - 0.29) | 0.09 (-0.91 - 0.57) |
| 2025-W39 | 0.05 (-0.47 - 0.34) | 0.03 (-0.36 - 0.29) | 0.09 (-0.85 - 0.57) |

Table S7: Comparison of estimates of vaccine impact against hospital admissions due to COVID-19 using 8-week and interpolated VE, October 2023 -– September 2024 and October 2024 -– September 2025, VEBIS-EHR, Europe.

| **Indicator** | |  | **Pooled** | **Belgium** | **Denmark** | **Italy** | **Navarre** | **Norway** | **Portugal** | **Sweden** |
| --- | --- | --- | --- | --- | --- | --- | --- | --- | --- | --- |
|  | **Hospital admission due to COVID-19** | | | | | | | | | |
| **65-79 years old** | |  |  |  |  |  |  |  |  |  |
| NAE (interpolated pooled VE) | | 2023-2024 | 2,542 (2,145 - 2,978) | 137 (104 - 173) | 684 (573 - 795) | 253 (211 - 296) | 27 (18 - 38) | 755 (643 - 875) | 121 (85 - 168) | 566 (470 - 670) |
|  | | 2024-2025 | 859 (686 - 1,033) | 48 (34 - 61) | 161 (129 - 196) | 32 (25 - 37) | 1 (-1 - 3) | 271 (216 - 331) | 8 (2 - 15) | 337 (270 - 404) |
| NAE (8-week pooled VE) | | 2023-2024 | 2,350  (2,038 – 2,692) | 125 (96 - 155) | 637 (568 - 714) | 234 (199 - 271) | 27 (18 - 37) | 696 (608 - 794) | 120 (83 - 162) | 513 (445 - 591) |
|  | | 2024-2025 | 777  (613 – 947) | 41 (28 - 54) | 151 (122 - 181) | 30 (25 - 35) | 1 (-1 - 3) | 244 (189 - 300) | 7 (1 - 13) | 305 (239 - 371) |
| **≥80 years old** | |  |  |  |  |  |  |  |  |  |
| NAE (interpolated pooled VE) | | 2023-2024 | 3,629 (3,089 - 4,432) | 149 (118 - 193) | 720 (619 - 861) | 526 (453 - 625) | 64 (51 - 85) | 698 (602 - 824) | 353 (268 - 470) | 1,119 (942 - 1,393) |
|  | | 2024-2025 | 1,315 (1,008 - 1,621) | 55 (38 - 71) | 192 (147 - 238) | 61 (46 - 72) | 1 (-2 - 3) | 272 (208 - 331) | 20 (7 - 34) | 716 (546 - 885) |
| NAE (8-week pooled VE) | | 2023-2024 | 3,112  (2,569 – 3,705) | 116 (76 - 157) | 636 (565 - 716) | 467 (368 - 565) | 45 (32 - 60) | 627 (528 - 740) | 255 (160 - 360) | 972 (820 - 1,156) |
|  | | 2024-2025 | 1,168  (795 – 1588) | 49 (29 - 71) | 170 (117 - 230) | 55 (36 - 73) | 0 (-2 - 3) | 248 (168 - 333) | 19 (4 - 36) | 626 (416 - 857) |

*NAE: number of averted events;
Values are shown as median (2.5th percentile, 97.5th percentile) across 1000 repetitions.*

Table S8: Comparison of estimates of vaccine impact against COVID-19-related deaths using 8-week and interpolated VE, October 2023 -– September 2024 and October 2024 –- September 2025, VEBIS-EHR, Europe.

| **Indicator** | |  | **Pooled** | **Belgium** | **Denmark** | **Italy** | **Navarre** | **Norway** | **Portugal** | **Sweden** |
| --- | --- | --- | --- | --- | --- | --- | --- | --- | --- | --- |
|  | **COVID-19-related death** | | | | | | | | | |
| **≥80 years old** | |  |  |  |  |  |  |  |  |  |
| NAE (interpolated pooled VE) | | 2023-2024 | 811 (559 - 1,028) | - | 188 (132 - 241) | 69 (45 - 85) | 7 (3 - 11) | 144 (104 - 175) | 208 (117 - 293) | 191 (134 - 246) |
|  | | 2024-2025 | 156 (98 - 208) | - | 27 (14 - 39) | 8 (5 - 10) | 0 (0 - 0) | 6 (2 - 8) | 32 (6 - 65) | 81 (49 - 115) |
| NAE (8-week pooled VE) | | 2023-2024 | 769  (520 – 1,040) | - | 187 (143 - 233) | 62 (32 - 91) | 7 (5 - 9) | 143 (114 - 171) | 185 (38 - 347) | 186 (135 - 239) |
|  | | 2024-2025 | 142  (77 – 212) | - | 24 (11 - 37) | 8 (4 - 11) | 0 (0 - 0) | 6 (4 - 8) | 28 (-17 - 63) | 78 (45 - 114) |

*NAE: number of averted events;
Values are shown as median (2.5th percentile, 97.5th percentile) across 1000 repetitions.*

### Figures

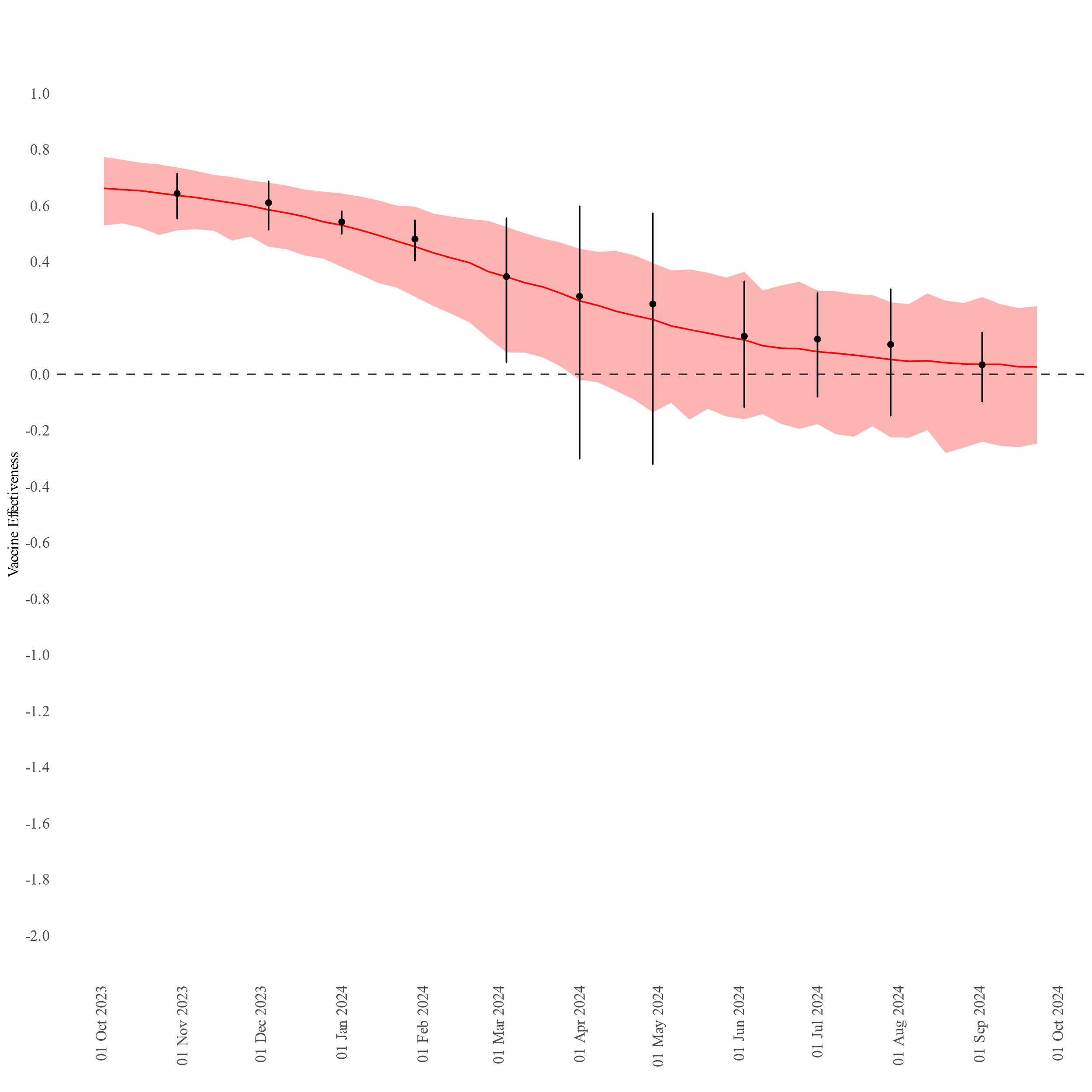

Figure S2: Weekly vaccine effectiveness estimates from the logistic decay interpolation model of the 8-week vaccine effectiveness estimates, hospital admission due to COVID-19, 65–79 years, 2023–2024, VEBIS-EHR, Europe. The line corresponds to the median weekly interpolated vaccine effectiveness and the ribbon contains the 95% prediction interval from 1000 samples. The points and error bars correspond to the observed 8-week vaccine effectiveness.

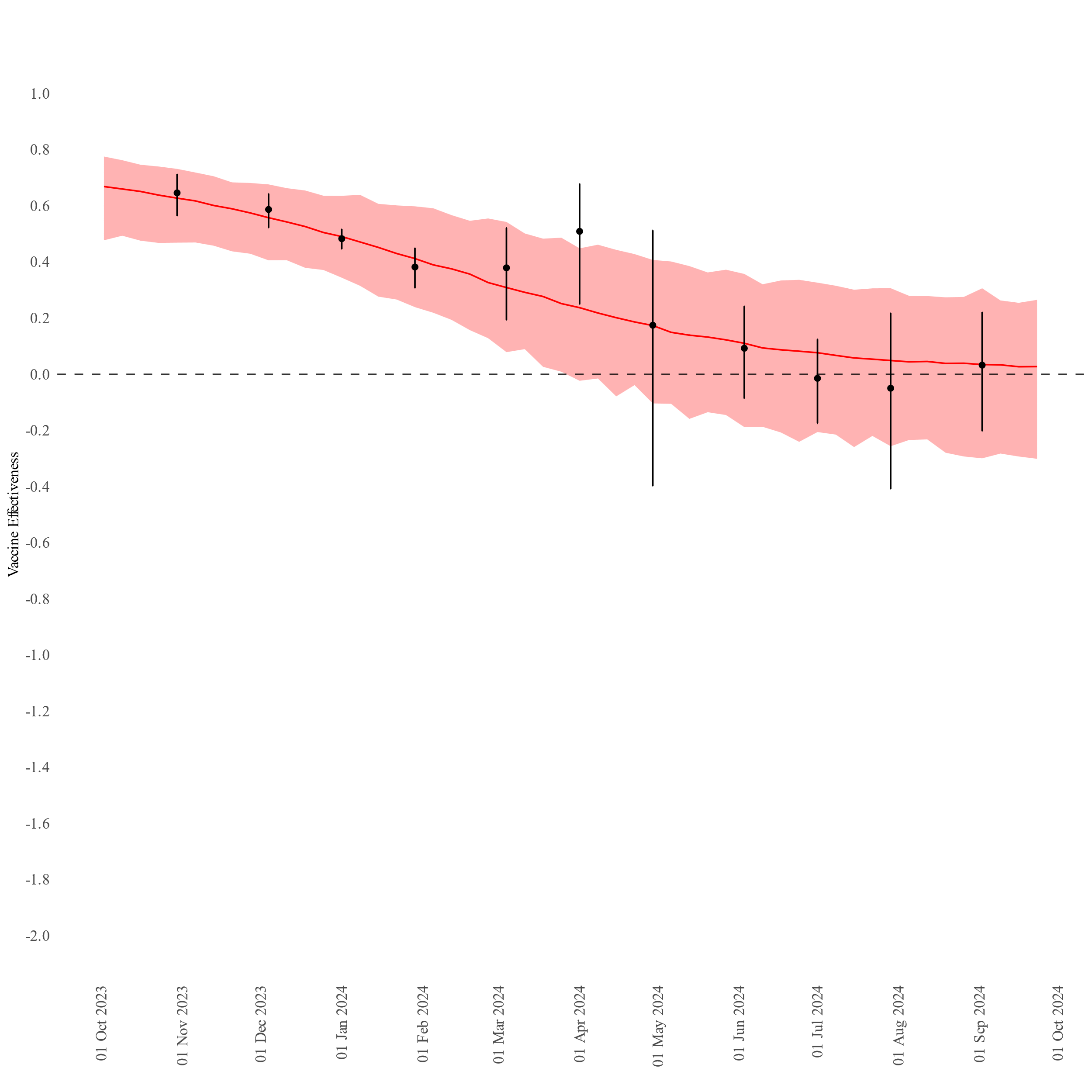

Figure S3: Weekly vaccine effectiveness estimates from the logistic decay interpolation model of the 8-week vaccine effectiveness estimates, hospital admission due to COVID-19, ≥80 years old, 2023–2024, VEBIS-EHR, Europe. The line corresponds to the median weekly interpolated vaccine effectiveness and the ribbon contains the 95% prediction interval from 1000 samples. The points and error bars correspond to the observed 8-week vaccine effectiveness.

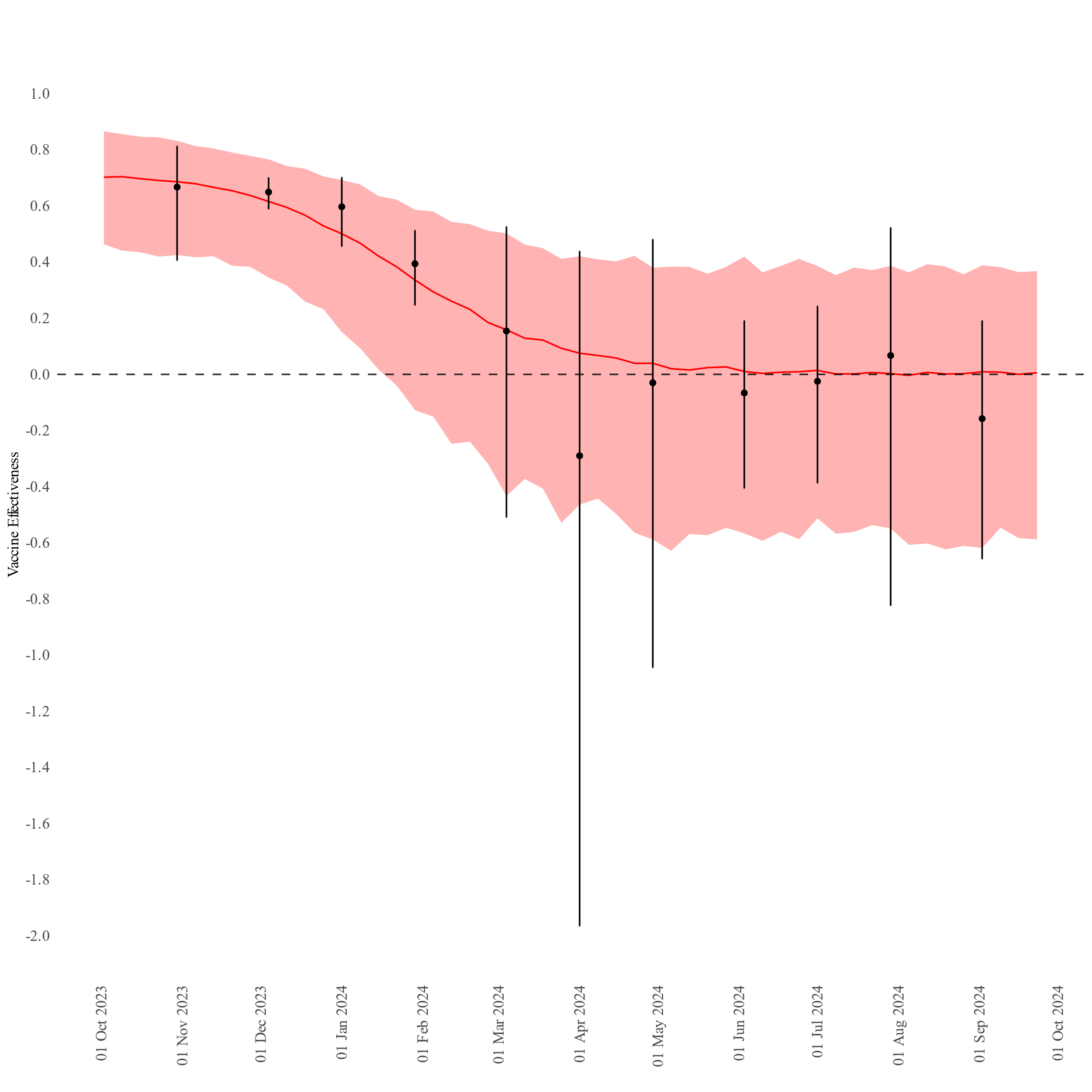

Figure S4: Weekly vaccine effectiveness estimates from the logistic decay interpolation model of the 8-week vaccine effectiveness estimates, COVID-19-related death, ≥80 years old, 2023–2024, VEBIS-EHR, Europe. The line corresponds to the median weekly interpolated vaccine effectiveness and the ribbon contains the 95% prediction interval from 1000 samples. The points and error bars correspond to the observed 8-week vaccine effectiveness.

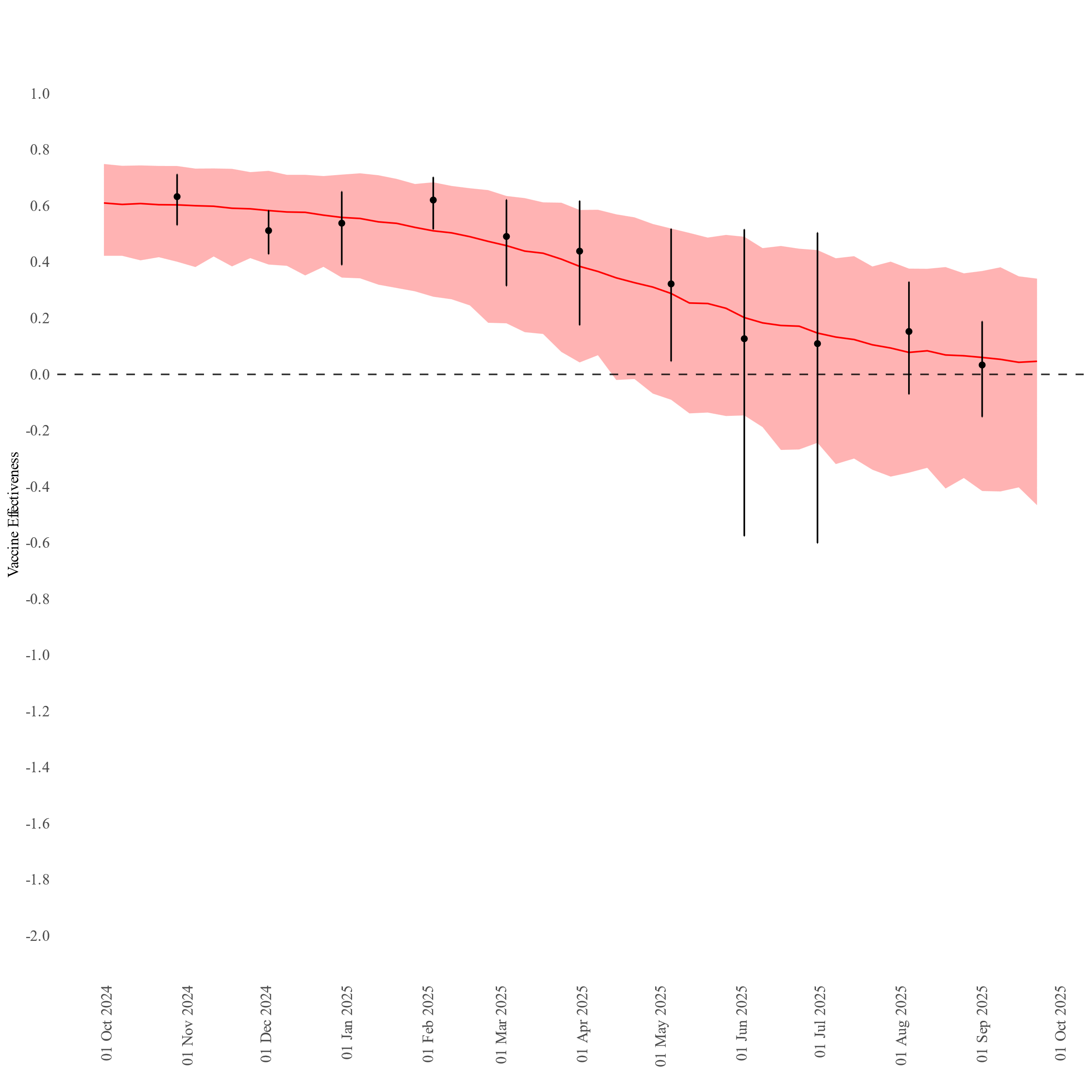

Figure S5: Weekly vaccine effectiveness estimates from the logistic decay interpolation model of the 8-week vaccine effectiveness estimates, hospital admission due to COVID-19, 65-79 years old, 2024-2025, VEBIS-EHR, Europe. The line corresponds to the median weekly interpolated vaccine effectiveness and the ribbon contains the 95% prediction interval from 1000 samples. The points and error bars correspond to the observed 8-week vaccine effectiveness.

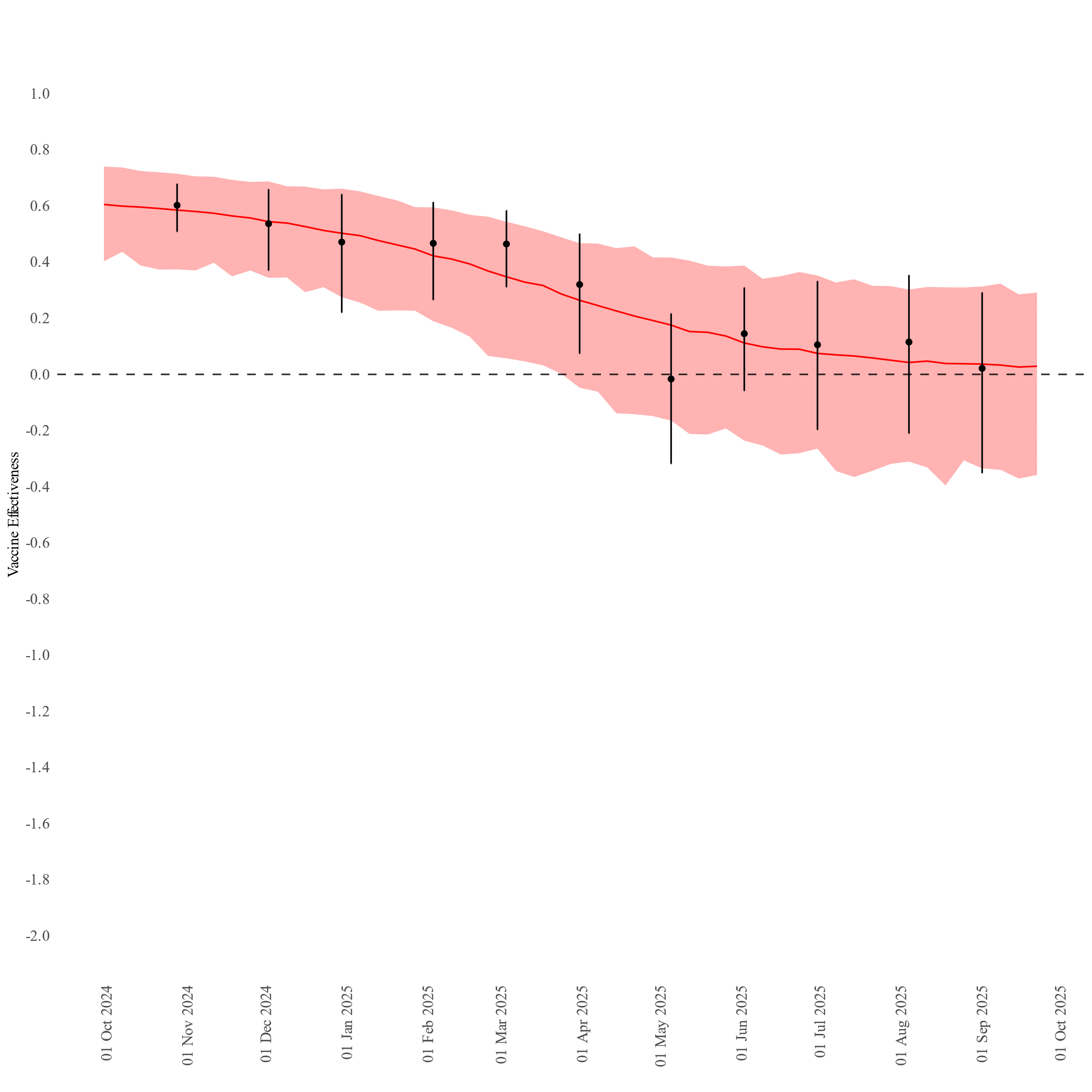

Figure S6: Weekly vaccine effectiveness estimates from the logistic decay interpolation model of the eight-week vaccine effectiveness estimates, hospital admission due to COVID-19, ≥80 years old, 2024–2025, VEBIS-EHR, Europe. The line corresponds to the median weekly interpolated vaccine effectiveness and the ribbon contains the 95% prediction interval from 1000 samples. The points and error bars correspond to the observed 8-week vaccine effectiveness.

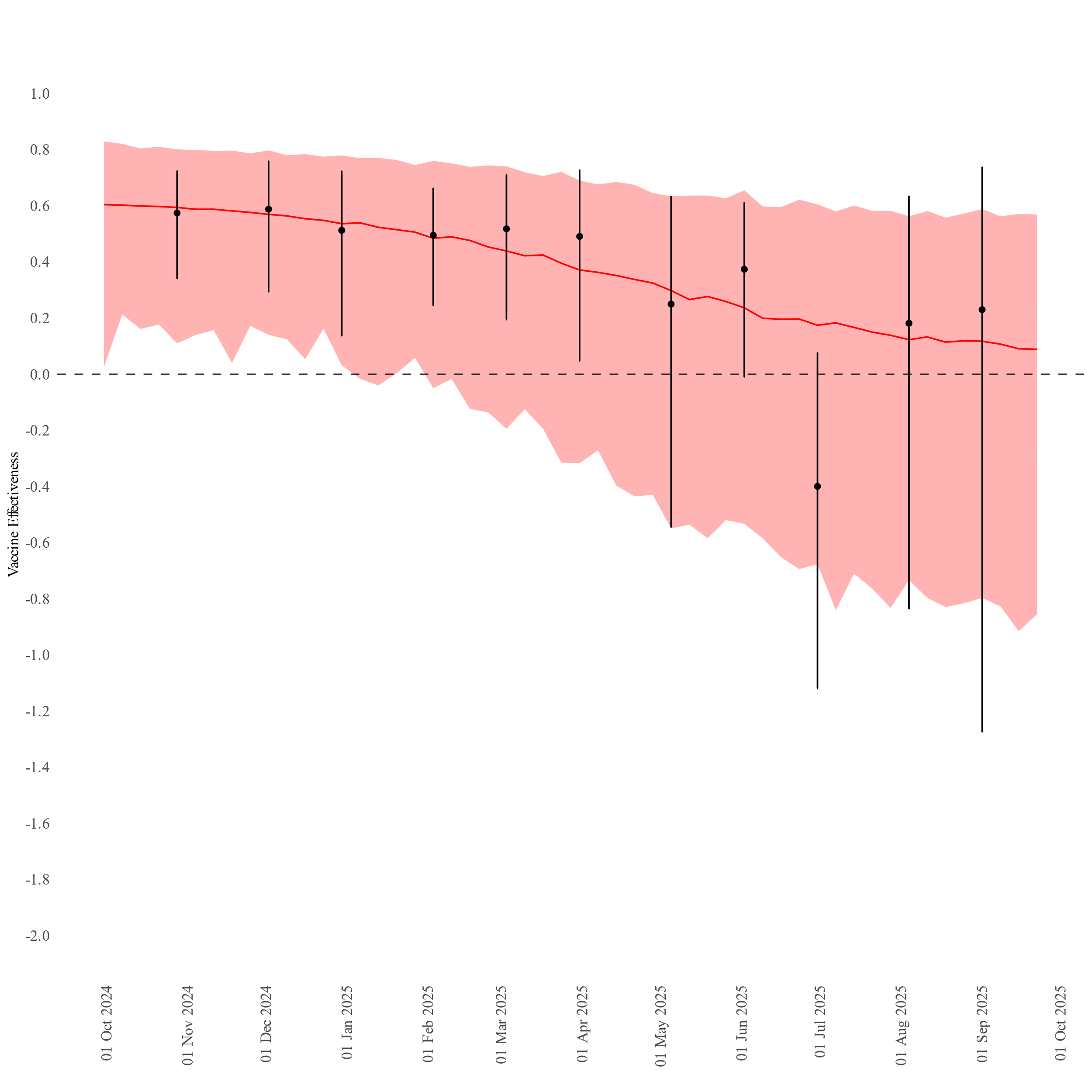

Figure S7: Weekly vaccine effectiveness estimates from the logistic decay interpolation model of the eight-week vaccine effectiveness estimates, COVID-19-related death, ≥80 years old, 2024–2025, VEBIS-EHR, Europe. The line corresponds to the median weekly interpolated vaccine effectiveness and the ribbon contains the 95% prediction interval from 1000 samples. The points and error bars correspond to the observed 8-week vaccine effectiveness.

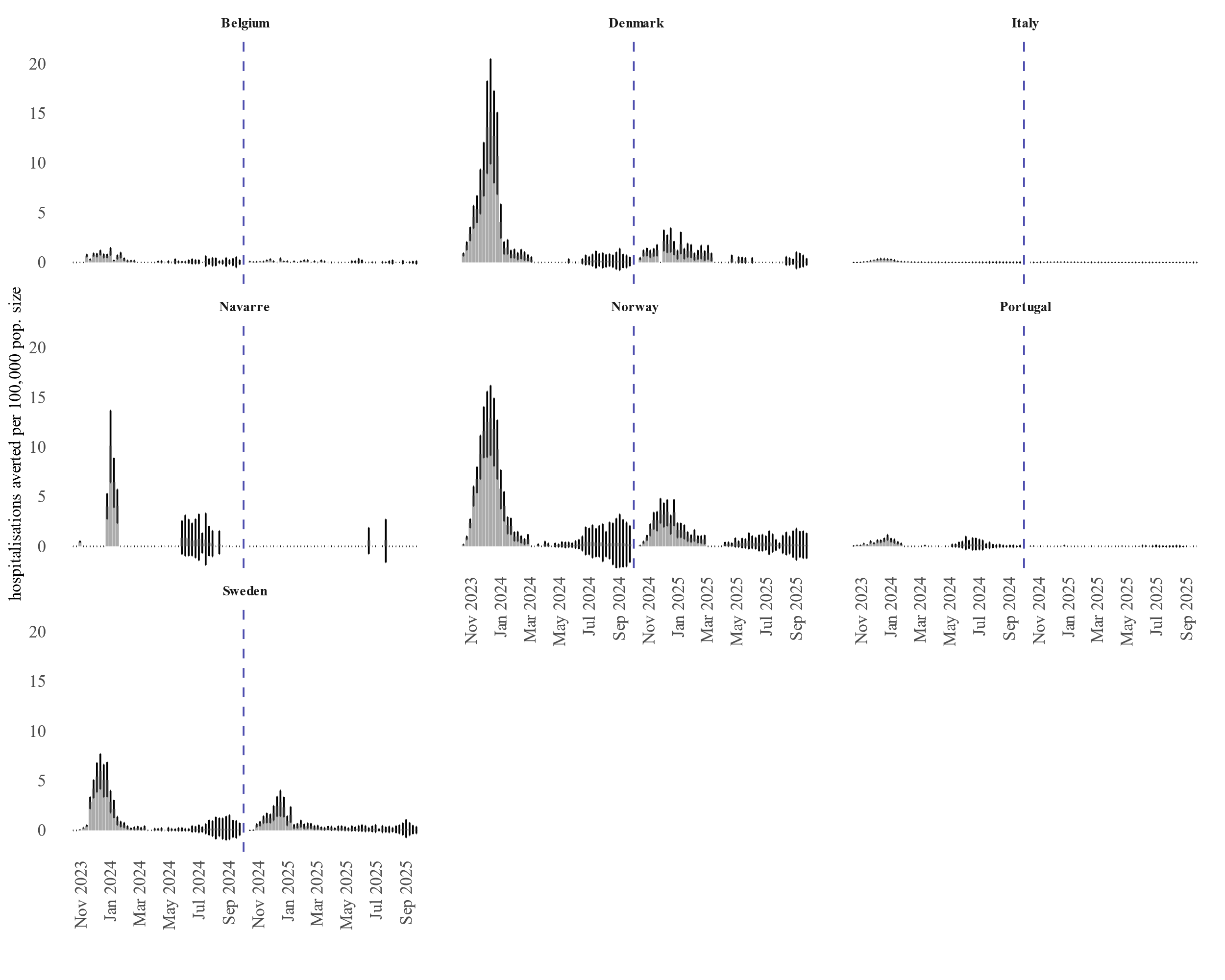

Figure S8: Weekly number of averted hospitalisations divided by the population size, 65–79 years, 2023–2024 and 2024-2025, VEBIS-EHR, Europe.

*Grey bars correspond to the median NAE divided by the population size; error bars correspond to 95% prediction intervals. Gaps correspond to weeks with no event observed.*

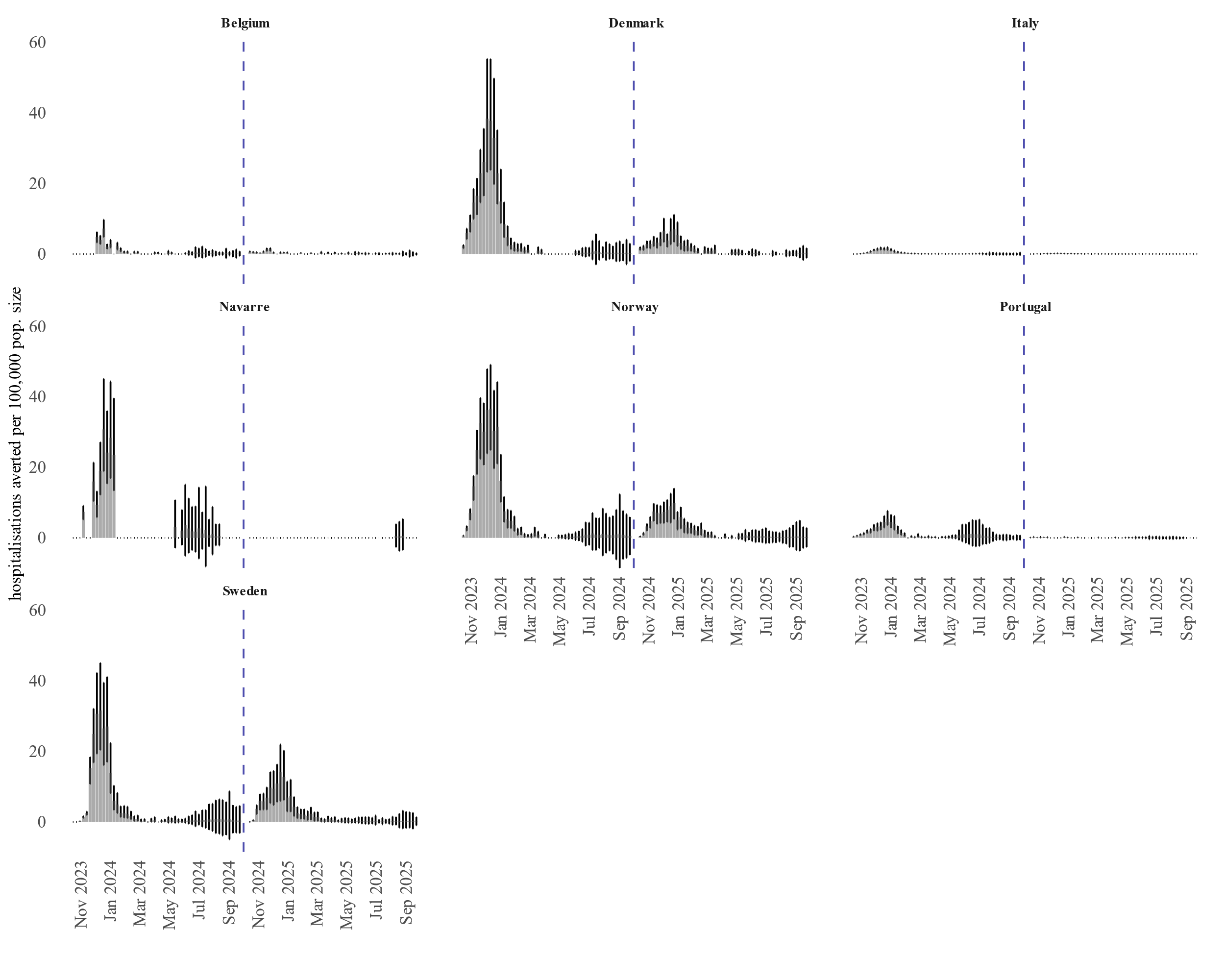

Figure S9: Weekly number of averted hospitalisations divided by the population size, ≥80 years old, 2023–2024 and 2024-2025, VEBIS-EHR, Europe.

*Grey bars correspond to the median NAE divided by the population size; error bars correspond to 95% prediction intervals. Gaps correspond to weeks with no event observed.*

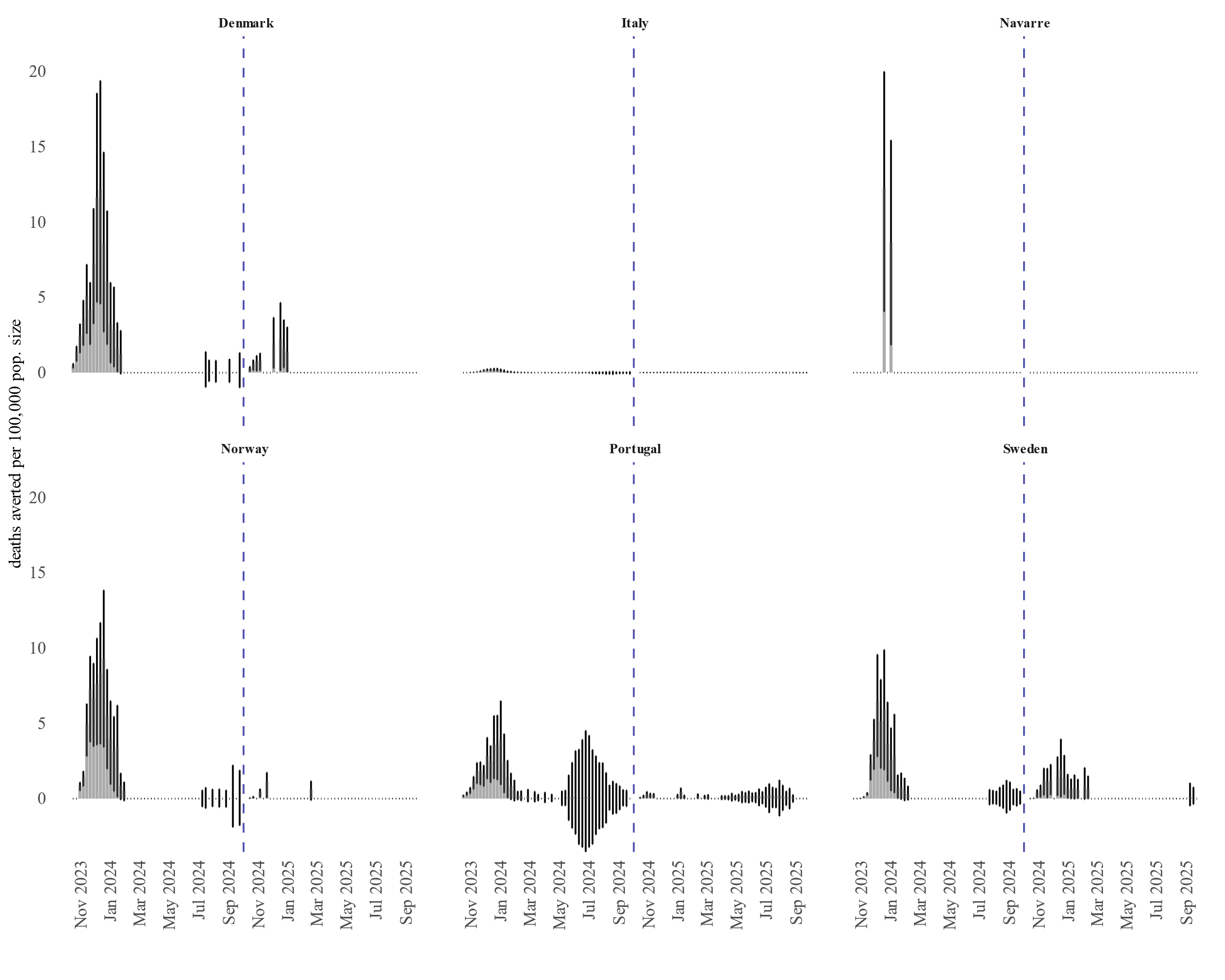

Figure S10: Weekly number of averted deaths divided by the population size, ≥80 years old, 2023–2024 and 2024-2025, VEBIS-EHR, Europe.

*Grey bars correspond to the median NAE divided by the population size; error bars correspond to 95% prediction intervals. Gaps correspond to weeks with no event observed.*
